# Differential COVID-19 Outcomes Across Lysosomal Disorders

**DOI:** 10.64898/2026.06.22.26356274

**Authors:** Blake K. Byer, Zachary Butzin-Dozier, Brenda M. McGrath, Joseph Muenzer, Lorne Clarke, Melissa A. Haendel, Shawn T. O’Neil

**Affiliations:** Department of Genetics, University of North Carolina at Chapel Hill, Chapel Hill, North Carolina, United States; Department of Pediatrics, Stanford University School of Medicine, Stanford, California, United States; OCHIN, Portland, Oregon, United States; Department of Pediatrics, University of North Carolina at Chapel Hill, Chapel Hill, North Carolina, United States; Department of Medical Genetics, University of British Columbia, Vancouver, British Columbia, Canada

**Keywords:** Lysosomal disorders, inborn errors of metabolism, electronic health records, computable phenotyping, COVID-19 severity, comorbidity burden, multiple correspondence analysis, retrospective cohort study

## Abstract

**Background:** Lysosomal disorders (LDs) are a heterogeneous group of rare inherited disorders characterized by multi-system involvement and high comorbidity burden, which raises concerns about severe COVID-19 outcomes. Conversely, because SARS-CoV-2 relies on endolysosomal pathways for cellular entry and replication, certain LDs may exert a protective effect against viral pathogenesis. Prior clinical evidence investigating LDs and severe SARS-CoV-2 infection has been limited by small sample sizes and inconsistent findings. Therefore, to resolve these conflicting biological hypotheses and estimate population-level outcomes, we conducted a large-scale retrospective cohort study using nationwide U.S. harmonized electronic health record data from the National Clinical Cohort Collaborative (N3C). This design utilized longitudinal records starting January 1, 2018, to evaluate COVID-19 infections captured between January 1, 2020, and July 11, 2024.

**Results:** The study included 16,380 individuals, comprising 5,460 patients with lysosomal disorders and 10,920 matched controls. Patients with LDs had significantly higher odds of COVID-19 hospitalization compared with controls (OR = 1.86, 95% CI: 1.70-2.04). Elevated odds were observed across the evaluated categories, but varied substantially. Notably, neurodegenerative LDs such as neuronal ceroid lipofuscinosis (OR = 9.32) and metachromatic leukodystrophy (OR = 2.33) remained associated with hospitalization after adjustment for comorbidities. Contrarily, the elevated odds for Fabry disease and Gaucher disease were no longer significant after adjustment. Mortality among hospitalized patients with LDs was comparable to that of matched controls (one-year survival: 82.1% vs 82.0%), suggesting that LD status does not independently worsen survival once hospitalization occurs.

**Conclusions:** Patients with LDs were at an increased odds of COVID-19 hospitalization, driven by a combination of elevated comorbidity burden and disorder-specific effects, which vary significantly across LD categories. This study clarifies that excess risk is concentrated in the transition to hospitalization. These patients may thus require personalized clinical care to mitigate the negative consequences of COVID-19.

## Background

Lysosomal disorders (LDs) are a heterogeneous group of rare inherited disorders caused by defects in lysosomal enzymes, transporters, or associated proteins, resulting in substrate accumulation, chronic inflammation, and progressive multisystem involvement [1]. Although individual LDs differ substantially in organ involvement, many share neurodegenerative, skeletal, renal, and cardiopulmonary phenotypes that mirror established risk factors for severe COVID-19.

Additionally, the lysosome itself is directly involved in SARS-CoV-2 pathogenesis. Viral entry occurs through the endosomal pathway, where the lysosomal protease Cathepsin L cleaves the spike protein to facilitate membrane fusion, and SARS-CoV-2 has also been shown to utilize lysosome-dependent pathways for egress [2–3]. Lysosomal dysfunction impairs autophagy, induces chronic inflammation, and causes airway cell death, all of which may exacerbate infection severity [4–5]. Conversely, certain LDs may alter the lysosomal environment, which may offer protection; for instance, Fabry disease may reduce susceptibility to SARS-CoV-2 by impairing ACE2 glycosylation and decreasing Cathepsin L activity [6].

Given that patients with LDs often have a high comorbidity burden, such as preexisting cardiopulmonary or renal conditions, there is concern that they would be particularly susceptible to severe outcomes during COVID-19 infection. However, the heterogeneous and complex alterations of cellular pathways across LDs may exacerbate or ameliorate viral pathogenesis. While some disorders can promote severe infection through chronic inflammation and impaired autophagy, others may disrupt the specific enzymatic and egress mechanisms required by the virus, thereby mitigating pathogenesis. The clinical impact of these competing mechanisms remains uncertain, necessitating a rigorous evaluation of severe COVID-19 outcomes, including hospitalization and mortality, among patients with LDs.

Current clinical evidence regarding these biological and clinical hypotheses remains limited and inconsistent, with findings drawn only from small observational studies. The CDC evaluated Gaucher disease in its review of conditions that may increase the risk of severe COVID-19; however, the association with hospitalization and mortality was deemed inconclusive due to small sample sizes, single-center studies, and the lack of appropriate control groups [7]. Fabry disease similarly shows mixed evidence. An international cohort of 22 Fabry disease patients suggested that most infections were mild to moderate, though two older patients with left ventricular hypertrophy and kidney disease experienced critical complications. However, findings on the association between Fabry disease and severe COVID-19 were inconclusive [8]. In a prospective single-center Fabry disease cohort, 13 of 104 patients developed SARS-CoV-2 infection, and only 2 of these 13 patients developed severe COVID-19; both were kidney transplant recipients on immunosuppression treatment. No patient died or was admitted to intensive care. The authors concluded that severe risk in Fabry disease may be driven mainly by immunosuppression in the context of kidney transplantation rather than Fabry disease itself [9].

For other LD types, the available literature is even sparser, mostly comprising single-center questionnaires and case reports. For instance, a single-center questionnaire study in Turkey examined 87 patients with LDs, including Gaucher, mucopolysaccharidosis (MPS), Fabry, and Pompe, and found no overall increase in COVID-19 risk compared to the general healthy population. Out of these 87 patients, only 8 tested positive for COVID-19, and two of these eight patients developed severe COVID-19 (one with MPS I and one with MPS IVA) [10]. Additionally, in a questionnaire sent to the Fabry, Gaucher, and Hunter Outcome registry sites, 80 of 91 (88%) respondents reported that the pandemic negatively affected standards of care, due to impaired communication and increased reliance on telemedicine, though there were minimal disruptions to intravenous enzyme replacement therapy (ERT) [11].

In cases where direct COVID-19 outcome data are lacking, several LDs have disorder-specific features that make severe respiratory infection biologically plausible. Gaucher disease, caused by glucocerebrosidase deficiency leading to glucocerebroside accumulation, is characterized by chronic immune dysregulation and systemic inflammation [12]. Fabry disease, resulting from globotriaosylceramide accumulation due to α-galactosidase A deficiency, is associated with progressive renal, cardiac, and vascular involvement [13]. In MPS, glycosaminoglycan deposition contributes to upper airway obstruction, recurrent respiratory infections, and restrictive lung disease, which may be exacerbated by kyphoscoliosis and pectus malformation [14]. LDs with neurodegeneration, such as metachromatic leukodystrophy and neuronal ceroid lipofuscinosis, commonly have dysphagia, which puts them at increased risk of aspiration pneumonia [15–16]. In cystinosis, a lysosomal cystine transporter disorder leading to cystine accumulation, advanced disease may involve respiratory muscle weakness and dysphagia, though these features are variable [17].

Taken together, the shared comorbidity burden, the heterogeneous and sometimes opposing roles of lysosomal pathways in viral pathogenesis, and the limited and inconsistent literature leave substantial uncertainty regarding COVID-19 risk among patients with lysosomal disorders. Because SARS-CoV-2 depends on endolysosomal machinery for cell entry, replication, and egress, a rigorous population-scale comparison of outcomes across biologically distinct LDs may inform which lysosomal perturbations exacerbate versus reduce viral pathogenesis, while simultaneously identifying which LD categories warrant intensified surveillance, earlier intervention, or tailored therapeutic strategies during COVID-19 or possibly other respiratory viral infections. Therefore, we conducted a large-scale retrospective cohort study using nationally sampled electronic health record data to evaluate the risk of SARS-CoV-2 infection and severe outcomes across multiple LD categories while accounting for demographic factors and comorbidities.

## Methods

### Data Source

This retrospective cohort study utilized data from the National Clinical Cohort Collaborative (N3C) COVID Enclave, a centralized resource and analytical platform for electronic health record (EHR) data in the United States. The enclave contains data from more than 22 million patients, including approximately 9 million COVID-19 cases and 14 million COVID-negative controls [18]. We used N3C data from version 180 (07/11/2024), which has 85 contributing sites and longitudinal data beginning January 1st, 2018. All ingested EHR data were standardized to the Observational and Medical Outcomes Partnership Common Data Model (OMOP CDM) to ensure consistency across sites [19].

### Lysosomal Disorder Patient Identification and Classification

Patients with LDs were identified using a Systematized Nomenclature of Medicine (SNOMED) concept codeset comprising 153 descendant terms of SNOMED:37155637 (“lysosomal storage disease”) and SNOMED:4053270 (“Disorder of lysosomal enzyme”), which identified 16,313 patients with LDs (full codeset in Supplementary Table 1).

To address the challenge of imprecise and varied temporal coding of LD diagnoses in the EHR, we developed an algorithm that leverages the hierarchical structure of SNOMED as a directed acyclic graph. Because LDs are inborn errors of metabolism, we used the patient’s entire record to identify them. For each patient, the algorithm selected the most specific LD diagnosis by selecting the maximally distant LD concept from the root nodes (SNOMED:37155637 or SNOMED:4053270). In the case of a tie in distance, the patient’s most recent diagnosis among the tied terms was selected. For instance, if a patient had sequential codes for “lysosomal storage disease” and “Fabry disease,” the algorithm assigned them to Fabry disease based on its greater distance from the root. This approach prioritized patients’ specific biological type, ensuring that they were classified by their most definitive diagnosis rather than being assigned to broad, nonspecific parent categories that often persist in clinical records.

For downstream regression analyses, we implemented a hierarchical roll-up strategy using the Mondo Disease Ontology (Mondo) [20]. Under this approach, patients assigned to specific (child) diagnoses were also included in all relevant higher-level (parent) disorder categories, resulting in non-mutually exclusive category groupings that reflect the ontology structure. Importantly, these assignments represent hierarchical relationships in which Mondo parent terms subsume corresponding SNOMED diagnoses, rather than exact cross-references or synonyms. For example, a patient with a SNOMED diagnosis of “Sanfilippo syndrome” (SCTID:4227905) would be rolled up into the broader “mucopolysaccharidosis” category (MONDO:0019249). The eleven Mondo parent categories used are: lysosomal lipid storage disorder (MONDO:0019245), sphingolipidosis (MONDO:0019255), Fabry disease (MONDO:0010526), mucopolysaccharidosis (MONDO:0019249), metachromatic leukodystrophy (MONDO:0018868), Gaucher disease (MONDO:0018150), neuronal ceroid lipofuscinosis (MONDO:0016295), gangliosidosis (MONDO:0017719), inborn disorder of lysosomal amino acid transport (MONDO:0019246), cystinosis (MONDO:0016239), and glycoproteinosis (MONDO:0017731). Full SNOMED-to-Mondo mappings are provided in Supplementary Table 1, LD category counts in Supplementary Tables 2 and 3, and the ontology hierarchy in Supplementary Figure 2.

### Key Definitions

#### COVID-19 Cases and Controls

COVID-19 cases were identified using a positive criterion from antigen or polymerase chain reaction (PCR) tests, or the International Classification of Diseases 10 Clinical Modification (ICD-10-CM) code U07.1 recorded between January 1st, 2020, and July 11, 2024, inclusive. The patient’s infection date was defined as the first of these criteria. Across both matched cohorts (below), controls were defined as patients without a documented lysosomal disorder.

#### Severity

COVID-19 severity was classified into five strata according to the World Health Organization Clinical Progression Scale [21–22]. The severity was assigned based on the highest documented severity: Mild No ED (WHO severity 1-3, outpatients with mild condition and no emergency department visit), Mild ED (WHO severity 3, outpatients with emergency department visit), Moderate (WHO severity 4-6, hospitalized patients without invasive ventilation), Severe (WHO severity 7-9, hospitalized patients with invasive ventilation or extracorporeal membrane oxygenation), and Death (WHO severity 10).

For multiple correspondence, regression, and survival analyses, severity was coded as Hospitalized (Moderate, Severe, and Death) or Non-Hospitalized (Mild No ED and Mild ED).

### Data Preprocessing and Cohort Construction

Patients with LDs were identified from N3C (v180, data through 11 July 2024) using a custom SNOMED codeset of 153 descendant terms. Prior to matching, demographic variables were standardized: missing or “Other” race was standardized to “Other/Unknown,” missing sex was set to “Unknown,” and invalid negative ages were replaced with zero. Missing values for age and body mass index (BMI) were imputed using predictive mean matching (PMM) with the *mice* package (version [3.17.0]) [23]. Imputation was performed separately for the LD and unmatched control arms to preserve their distinct demographic distributions.

To minimize confounding by indication, demographic, and geographic bias, we constructed two 1:2 Mahalanobis distance matched cohorts using the *MatchIt* package (version [4.7.2]) [24]. Matching was performed using Mahalanobis distance on age, race, sex, and the natural logarithm of days observed (calculated pre-COVID for the COVID cohort), with exact matching on the data-contributing site. The COVID Matched Cohort (N = 16,380) restricted both arms to positive COVID-19 cases with at least one healthcare encounter prior to the index date. This cohort comprised 5,460 patients with LDs and 10,920 controls, and was used to evaluate acute phenotypes, clinical severity, and mortality (Figure 1). The Baseline Matched Cohort (N = 48,357) restricted both arms to at least two healthcare encounters and matched patients regardless of COVID-19 status. This cohort comprised 16,119 patients with LDs and 32,238 controls, and was used to model baseline infection risk (Supplementary Figure 1).

**Figure 1:**
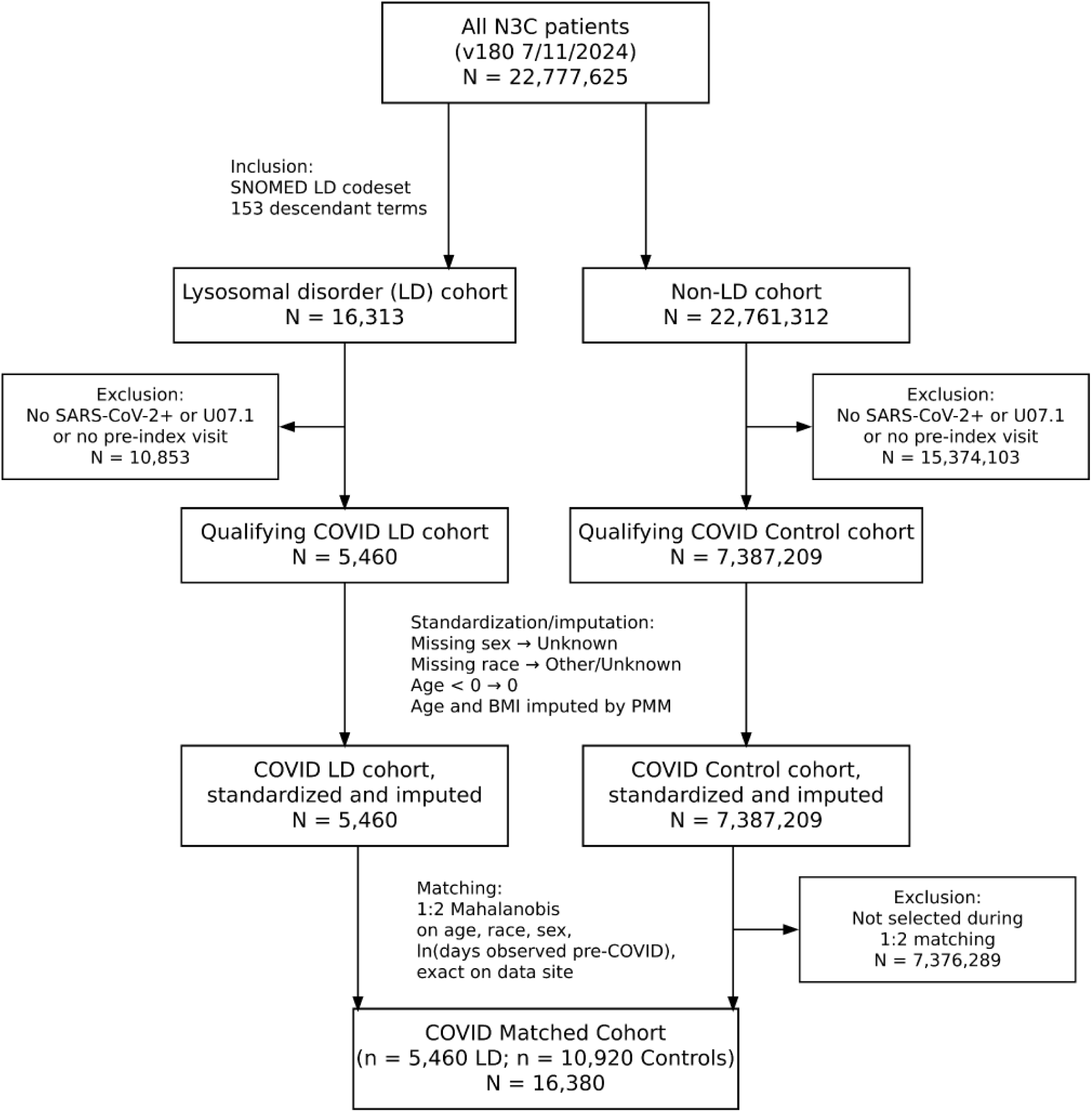
Cohort construction of the COVID Matched Cohort. Flowchart of inclusion and exclusion criteria applied to the N3C population. Standardization, imputation, and 1:2 Mahalanobis matching were performed to construct a cohort comprising 5,460 patients with LDs and 10,920 controls.

### Comorbidity Assessment

Comorbidity burden was assessed using 31 Elixhauser comorbidities derived from patients’ ICD codes, as described by Quan et al. [25]. The Elixhauser index was selected over the Charlson index because it includes 14 additional hospitalization predictors and has demonstrated superior performance at predicting in-hospital mortality when using the weights developed by Sharma et al. [26].

Additionally, we calculated an Elixhauser comorbidity score for each patient based on these weights. Mappings and score calculations were performed with the *comorbidity* package (version [1.1.0]) [27]. To prevent artificial score inflation due to double-counting, only the most severe instance was recorded for conditions with both complicated and uncomplicated forms (e.g., hypertension). While there aren’t canonical organ system groups in the Elixhauser mapping, we classified the 31 comorbidities into 13 organ system and disease categories for tabulation of prevalence summaries (Supplementary Table 8).

### Statistical Analysis

All statistical modeling was conducted within the N3C enclave using SQL and R (4.3.3). Statistical significance was defined for p-values < 0.05 unless otherwise noted, and 95% confidence intervals for all estimated odds and hazard ratios are reported. For disorder category-specific analyses, Benjamini-Hochberg false discovery rate (FDR) correction was applied which are reported as FDR q-values. This study was reported in accordance with the STROBE statement guidelines, utilizing the RECORD extension for routinely collected health data; the completed checklist is provided in the Supplementary Material [28–29].

#### Weighted multiple correspondence analysis

To characterize the latent structure among categorical comorbidities and to stratify LD cases and controls in the COVID Matched Cohort, we performed weighted multiple correspondence analysis (MCA) using the *FactoMineR* package, a multivariate dimensionality-reduction method for categorical data (version [2.12]) [30].

The MCA was performed on an indicator matrix with 65 categorical levels, derived from 31 binary Elixhauser comorbidities (recorded at any time relative to COVID) and a three-level healthcare utilization metric (low, moderate, high; assigned by the 25th, 50th, and 75th percentiles of annual visits). Demographic variables were excluded since they were already balanced in the matching procedure. Controls were assigned a weight of 0.5 to balance group contributions.

Group separation was assessed using Multivariate Analysis of Variance (MANOVA) on the coordinates of the first 20 dimensions (explaining 78.2% of the total inertia). Cohort status (LD vs control), clinical trajectory (hospitalized vs non-hospitalized), and their interaction were treated as supplementary variables that did not contribute to axis construction. The overall significance of group separation across these categories was determined using Pillai’s Trace. We then performed post hoc univariate Tukey’s Honest Significant Differences (HSD) tests on the scores for each individual dimension to identify dimensions driving group differences and their direction.

#### Conditional logistic regression

We used conditional logistic regression to estimate the odds of SARS-CoV-2 infection in the Baseline Matched Cohort and severe outcomes (hospitalization and mortality) in the COVID Matched Cohort. To account for the matching structure, each model was conditioned on the matching subclass, which effectively adjusted for age, race, sex, data-contributing site, and the natural logarithm of days observed by comparing patients with LDs directly to their specific matched controls. For the Baseline Matched Cohort, we fitted models for 10 LD categories and 1 overall LD group (11 models in total). For the COVID Matched Cohort, where data were more limited, we fitted models for 8 LD categories plus one overall LD group (9 models total).

To assess the contributions of comorbidities to LD-associated risk, we identified Elixhauser comorbidities that differed in prevalence between LD categories and matched controls prior to or on the COVID index date using logistic regression. Comorbidities present in at least five patients and meeting p < 0.001 were included as covariates in adjusted conditional logistic regression models.

#### Survival analysis

Survival probabilities following COVID-19 infection were assessed using the Kaplan-Meier survival estimator from the *ggsurvfit* package (version [1.1.0]) [31]. Survival analysis was performed over an 1800-day (∼5-year) follow-up period post-COVID. The two-sided log-rank test was used to determine differences in survival curves. We also determined differences in median time-to-death post-COVID between the hospitalized and non-hospitalized groups, and between LD and COVID control patients, using a two-sided Wilcoxon rank-sum test.

Finally, we evaluated the impact of acute-phase treatments (procedures or drugs administered in the first 14 days after the COVID index date) using Cox proportional hazards models from the *survival* package (version [3.8_3]). [32]. The model assessed survival following administration of 15 distinct drugs and 11 distinct standard-of-care COVID-19 procedures and treatments (full codesets are available in N3C; codeset IDs listed in Supplementary Tables 4 and 5). To prevent immortal time bias, the landmark date was set at 15 days post-COVID, and only patients who died on or after that date were included in the survival analysis. All Cox models were adjusted for the patient’s Elixhauser score, where missing scores were replaced with zero, and conditioned on the matching subclass. Additionally, we tested for interaction effects between LD status and the administration of these interventions.

## Results

From N3C, which contained 22,777,625 patients as of July 11, 2024, we identified a primary population of 16,313 patients with LDs. Our inclusion criteria required at least two healthcare encounters for the Baseline Matched Cohort (N = 16,119 LD) and at least one encounter prior to the infection index date (between January 1st, 2020, and July 11, 2024, inclusive) for the COVID Matched Cohort (N = 5,460 LD).

### Matched cohort characteristics

To minimize confounding by indication, demographic, and geographic bias, we constructed two 1:2 Mahalanobis distance matched cohorts. First, a Baseline Matched Cohort (N = 48,357) matched LD cases to COVID-agnostic controls and was used to model infection risk and to characterize LD phenotypes (Supplementary Figure 1). Additional information on the Baseline Matched Cohort is provided in Supplementary Tables 6 and 7. Second, a COVID Matched Cohort (N = 16,380) matched COVID-positive LD to COVID-positive controls to evaluate acute phenotypes, clinical severity, and mortality (Figure 1). Both cohorts utilized exact matching on data-contributing site and showed high demographic parity across all matching variables (SMD < 0.1; Supplementary Figures 3 and 4).

The COVID Matched Cohort was well-balanced across all primary demographic variables, with a median age of 54 (IQR: 34, 68) and a female majority (56%). The racial distribution was majority White (78%), followed by Black or African American (9.9%) and Asian (2.6%). Before imputation, in LDs, age was 4.6% missing, and BMI was 14.3% missing; in controls, age was 4.6% missing, and BMI was 39% missing. Both groups were observed for a median of 1,315 days prior to COVID, providing sufficient longitudinal data for comorbidity assessment. Despite nearly identical demographics, patients with LDs utilized healthcare at significantly higher rates, with a median of 61 pre-COVID visits compared to 31 in controls. Vaccination status was comparable between groups, with the majority (70%) of patients lacking a documented COVID-19 vaccination (Table 1).

**Table 1:** Characteristics of the COVID Matched Cohort. Summary table for patients in the COVID Matched Cohort, including severity, healthcare utilization, vaccination, clinical factors, demographics (age, race, sex), and geographic region. Discrete data are displayed as “count (percentage)” and continuous data as “median (IQR)”.

| Characteristic | Control<br>N = 10,920 | LD<br>N = 5,460 |
| --- | --- | --- |
| **Severity Type** |  |  |
| Mild No-ED | 7,868 (72%) | 3,425 (63%) |
| Mild ED | 1,584 (15%) | 864 (16%) |
| Moderate | 1,233 (11%) | 967 (18%) |
| Severe | 51 (0.5%) | 78 (1.4%) |
| Death | 184 (1.7%) | 126 (2.3%) |
| **Median Observation Days (IQR)** |  |  |
| Before COVID | 1,315 (973, 1,591) | 1,315 (971, 1,591) |
| Post COVID | 661 (288, 965) | 759 (404, 1,031) |
| **Healthcare Utilization, Median Visit Count (IQR)** |  |  |
| Visits Before COVID | 31 (11, 74) | 61 (24, 129) |
| Visits Post COVID | 20 (6, 50) | 41 (16, 92) |
| **Vaccination (Doses Before or on COVID)** |  |  |
| 0 | 7,599 (70%) | 3,827 (70%) |
| 1 | 502 (4.6%) | 271 (5.0%) |
| 2 | 1,332 (12%) | 646 (12%) |
| 3 | 974 (8.9%) | 467 (8.6%) |
| 4 | 513 (4.7%) | 249 (4.6%) |
| **Clinical Factors** |  |  |
| Long COVID | 220 (2.0%) | 153 (2.8%) |
| Reinfection | 748 (6.8%) | 576 (11%) |
| BMI | 30 (26, 36) | 31 (26, 37) |
| **Race** |  |  |
| White | 8,566 (78%) | 4,283 (78%) |
| Black or African American | 1,076 (9.9%) | 538 (9.9%) |
| Asian | 288 (2.6%) | 144 (2.6%) |
| American Indian or Alaska Native | 46 (0.4%) | 23 (0.4%) |
| Other or Unknown | 944 (8.6%) | 472 (8.6%) |
| **Sex** |  |  |
| Female | 6,144 (56%) | 3,071 (56%) |
| Male | 4,474 (41%) | 2,237 (41%) |
| Unknown | 302 (2.8%) | 152 (2.8%) |
| Age | 54 (34, 68) | 54 (34, 68) |
| Elixhauser Score | 2 (-2, 13) | 12 (0, 29) |
| **Region** |  |  |
| Atlantic | 2,604 (24%) | 1,197 (22%) |
| Central | 5,807 (53%) | 3,009 (55%) |
| Southern | 793 (7.3%) | 402 (7.4%) |
| Western-Pacific | 1,082 (9.9%) | 520 (9.5%) |
| Unknown | 634 (5.8%) | 332 (6.1%) |

### Comorbidity stratification between LD and matched controls

In the COVID Matched Cohort, patients with LDs exhibited a substantially higher baseline comorbidity burden, with a median Elixhauser score of 12 (IQR: 0, 29) compared to 2 (IQR: -2, 13; p < 0.001) for COVID controls.

In the MCA analysis of comorbidity burden, the first two dimensions explained 26.3% of the inertia. Dimension 1 (19.9% of inertia) represented a cardiovascular-utilization-comorbidity burden gradient, evident by an Elixhauser score gradient along Dim. 1 (Figure 2b), driven by congestive heart failure (contribution = 5.4%, η² = 0.44), complicated hypertension (5.3%, η² = 0.45), and pulmonary circulation disorders (4.6%, η² = 0.35, Figure 2b,c). Dimension 2 accounted for an additional 6.3% of the inertia and was shaped by alcohol use (10%, η² = 0.28), complicated hypertension (9.4%, η² = 0.25), and renal failure (7.9%, η² = 0.21, Figure 2d). These findings align with the progressive multi-system dysfunction, including cardiac, renal, and pulmonary involvement characteristic of lysosomal defects.

**Figure 2:**
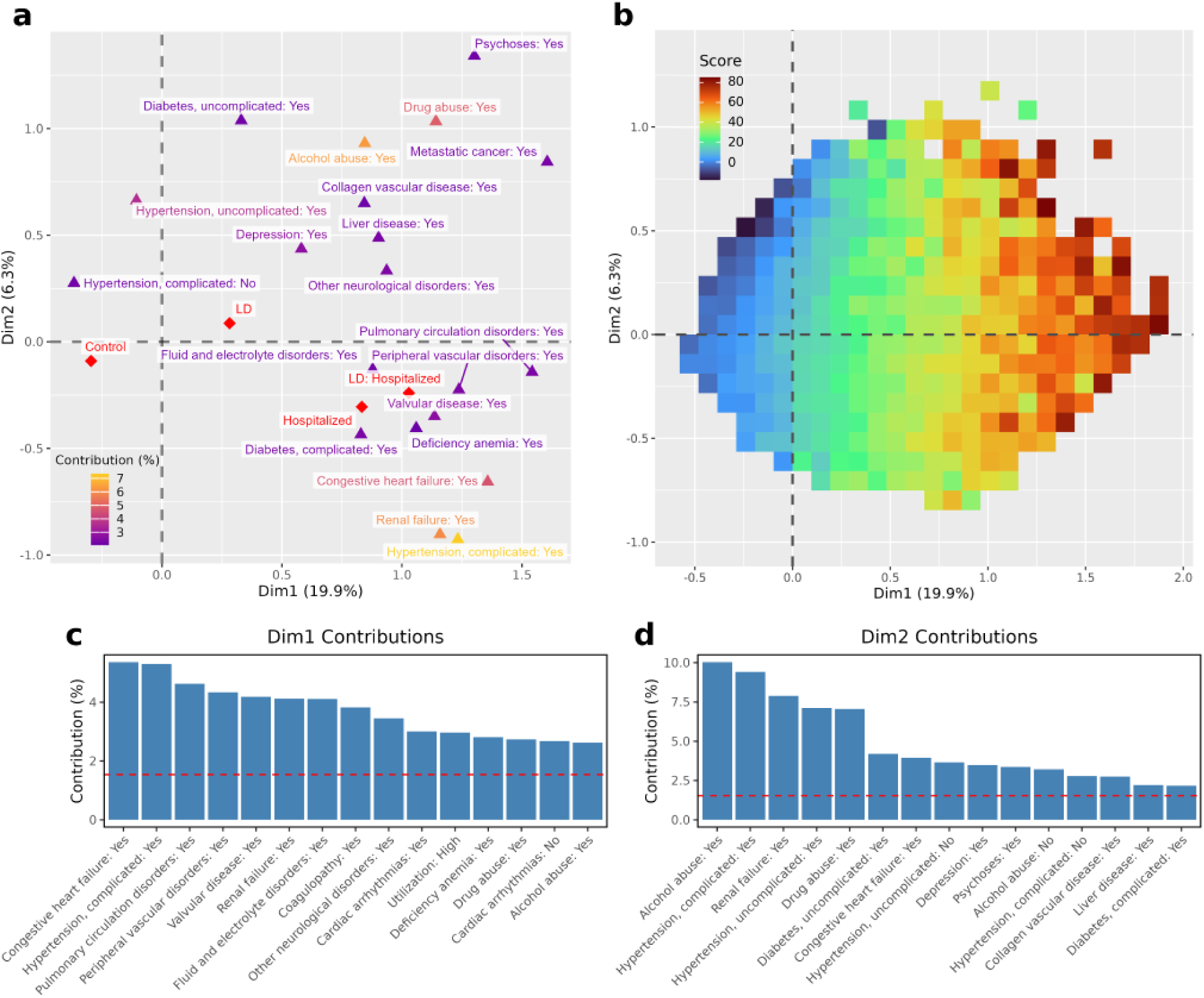
Multiple correspondence analysis differentiates LD and control patients in comorbidity latent space. **a,** Top twenty comorbidity variables ranked by average percentage contribution in dimensions 1 and 2, plotted in MCA space colored by contribution. Points represent the centroids of patients with each condition, with supplementary points (red) projected onto the MCA space. **b,** Binned heatmap of the MCA space, with each cell colored by the median patient Elixhauser comorbidity score. **c, d,** Top 15 contributing variables to dimension 1 (Dim1, **c**) and dimension 2 (Dim2, **d**). In both panels, the red horizontal dashed line indicates the expected contribution if all 65 variables contributed equally (1/65 ≈ 1.5%).

We used MANOVA and post hoc univariate tests to determine whether these latent dimensions could distinguish between groups. An analysis across the first 20 dimensions (explaining 78.2% of the total inertia) showed significant separation between LD and control patients (Pillai’s Trace = 0.113, F(20,15,656) = 99.46, p < 0.001) and between COVID-19 hospitalized and non-hospitalized patients (Pillai’s Trace = 0.210, F(20, 15,656) = 208.68, p < 0.001). Dimension 1 was the primary driver of separation for both contrasts: patients with LDs were significantly shifted toward higher Dim. 1 scores relative to controls (η² = 0.083, F(1, 15,675) = 1397.7; Tukey mean difference = 0.262, p < 0.001), and hospitalized patients were similarly shifted relative to non-hospitalized patients (η² = 0.146, F(1, 15,675) = 2723.6; Tukey mean difference = 0.455, p < 0.001). Additional dimensions showed significant group differences, but with considerably smaller effect sizes (η² < 0.04).

Supplementary variables projected onto the MCA space in Figure 2a show that both LD and hospitalized patients shift toward the high-burden right half-plane; the LD-hospitalized centroid was pulled toward peripheral vascular disorders, pulmonary circulation disorders, and valvular disease. This suggests that LD status creates a specific vascular-pulmonary phenotype that exacerbates the standard COVID-19 hospitalization signature. Patient distributions are shown in more detail in Supplementary Figure 5.

These latent patterns were reflected in the observed prevalence of comorbidities, and unique patterns emerged across severity strata. Although patients with LDs had a greater overall comorbidity burden, the differences were most pronounced in the Mild ED group (mean Δ = 11.9%) and narrowed among non-hospitalized patients (Mild No ED, mean Δ = 7.4%; Supplementary Table 9). Hierarchical clustering of these prevalence differences in Figure 3a identified phenotypic “hot” and “cold” zones: patients with LDs in the Mild ED group showed higher cardiac, substance use, and neurologic deficits than controls, yet Severe patients with LDs had reduced cardiometabolic deficits, such as congestive heart failure and hypertension. Mean prevalence of comorbidities across major organ systems showed that patients with LDs had higher prevalence earlier in the clinical severity spectrum than controls, with the strongest trends for pulmonary and nutritional conditions (Figure 3b).

**Figure 3:**
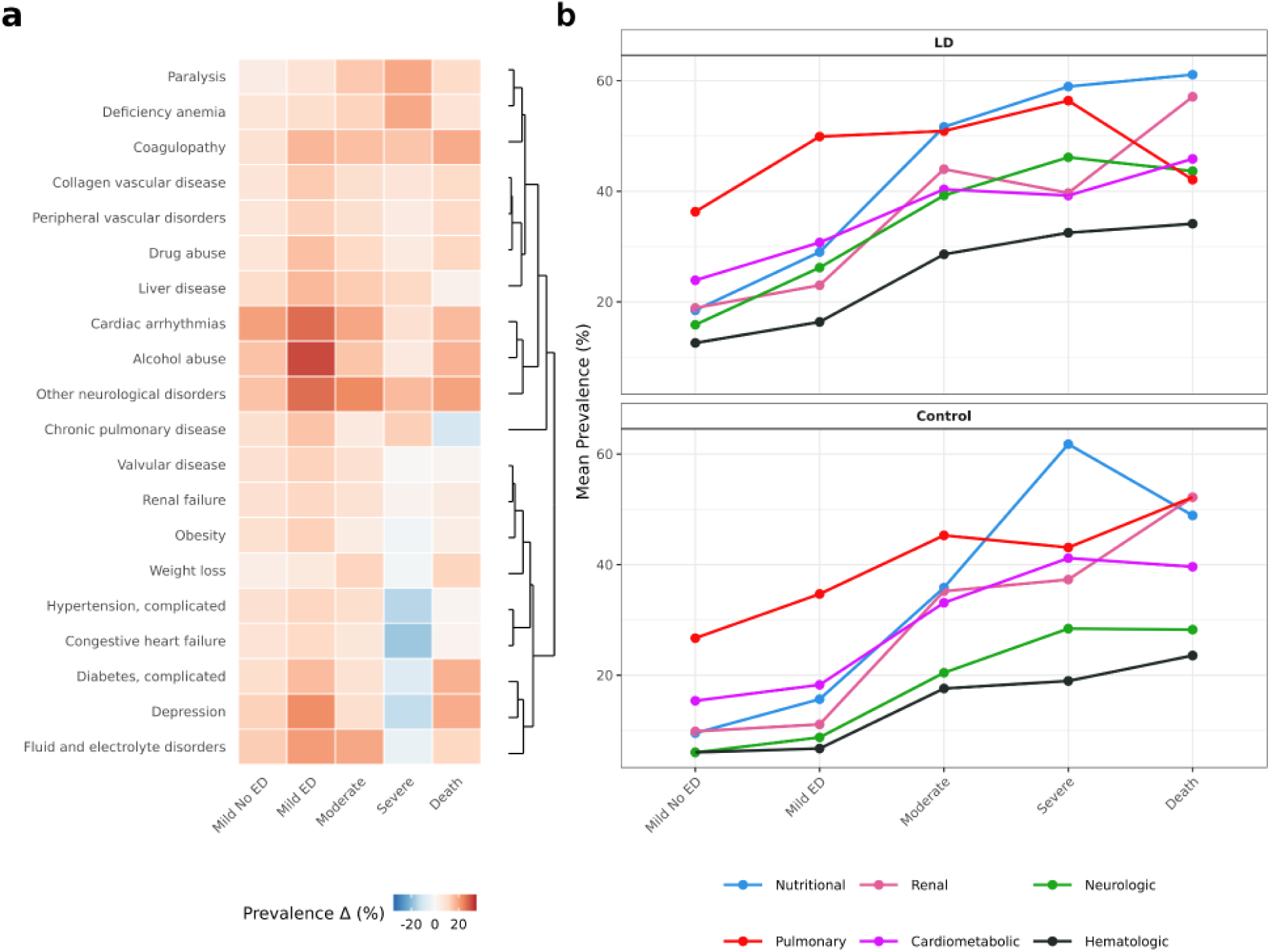
Increased comorbidity burden in patients with LDs vs controls at milder clinical severity strata. **a,** Elixhauser comorbidity prevalence difference (Δ). Heatmap displaying the percentage point difference in prevalence between patients with LDs (n = 5,384) and controls (n = 10,293) across increasing COVID-19 severity strata. Positive values (red) indicate higher prevalence in the LD cohort, while negative values (blue) indicate higher prevalence in controls. The top 20 comorbidities, ranked by the maximum prevalence difference, are shown. The dendrogram represents the hierarchical clustering of these differences, grouping comorbidities with similar enrichment patterns across the clinical severity spectrum. **b,** Organ system comorbidity prevalence. Mean comorbidity prevalence for the top six organ system categories, selected based on the maximum difference between Death and Mild No ED strata. Profiles are stratified by cohort (LD top, control bottom) and plotted across the five clinical severity strata (increasing from left to right).

### LD and matched controls have no difference in odds of SARS-CoV-2 infection

In the Baseline Matched Cohort, there were no significant differences in the odds of SARS-CoV-2 infection between patients with LDs and control patients (OR = 0.98, 95% CI: 0.94-1.02, p = 0.287; Supplementary Table 10). There were also no significant differences in the ten LD categories (Supplementary Table 10). This null baseline ensures that subsequent changes in hospitalization odds (below) are not driven by differential infection odds.

### Conditional logistic regression indicates LD categories’ odds for severe COVID

To evaluate the association between LD categories and severe outcomes (hospitalization), we used conditional logistic regression, with separate models for each LD category.

In logistic regression models conditioned on the matching subclass, patients with lysosomal disorders had significantly higher odds of COVID-19 hospitalization than matched controls (OR = 1.86, 95% CI: 1.70-2.04; Table 2). Elevated odds were observed across all estimable LD categories, with particularly high odds for neuronal ceroid lipofuscinoses (OR = 15.18, 95% CI: 5.36-42.98) and metachromatic leukodystrophy (OR = 3.50, 95% CI: 2.10-5.86).

**Table 2:** Conditional logistic regression for hospitalization in the COVID Matched Cohort by lysosomal disorder category. Odds ratios (OR) for severe COVID-19 outcomes. Models were conditioned on matching variables (age, sex, race, days observed, and data-contributing site). Adjusted models additionally included comorbidity covariates (Supplementary Table 11) and were estimated only for Mondo LD categories. “—” indicates not estimated. Mucopolysaccharidosis was not estimated due to dataset limitations; inborn disorder of lysosomal amino acid transport was not estimated because it completely overlapped with cystinosis. “†” indicates no adjusted model was estimated because there were no significant covariates.

|  | Unadjusted Model |  |  | Covariate-Adjusted Model |  |  |
| --- | --- | --- | --- | --- | --- | --- |
| LD Category | OR (95% CI) | p-value | FDR q-value | OR (95% CI) | p-value | FDR q-value |
| Lysosomal disorder (Overall) | 1.86 (1.70, 2.04) | < 0.001 | < 0.001 | — | — | — |
| Sphingolipidosis | 2.10 (1.84, 2.39) | < 0.001 | < 0.001 | 1.43 (1.19, 1.71) | < 0.001 | < 0.001 |
| Lysosomal lipid storage disorder | 1.77 (1.61, 1.95) | < 0.001 | < 0.001 | 1.14 (1.02, 1.28) | 0.023 | 0.032 |
| Fabry disease | 1.81 (1.24, 2.62) | 0.002 | 0.002 | 1.02 (0.64, 1.62) | 0.948 | 0.948 |
| Gaucher disease | 1.90 (1.01, 3.58) | 0.047 | 0.047 | 1.57 (0.79, 3.12) | 0.194 | 0.226 |
| Metachromatic leukodystrophy | 3.50 (2.10, 5.86) | < 0.001 | < 0.001 | 2.33 (1.28, 4.27) | 0.006 | 0.014 |
| Gangliosidosis | 3.71 (1.65, 8.31) | 0.001 | 0.002 | 3.50 (1.37, 8.95) | 0.009 | 0.015 |
| Cystinosis | 5.03 (2.11, 11.98) | < 0.001 | < 0.001 | † | † | † |
| Neuronal ceroid lipofuscinosis | 15.18 (5.36, 42.98) | < 0.001 | < 0.001 | 9.32 (2.91, 29.84) | < 0.001 | < 0.001 |

After adjustment for comorbidity covariates, effect estimates were attenuated but still statistically significant for several categories, including sphingolipidosis, lysosomal lipid storage disorder, metachromatic leukodystrophy, gangliosidosis, and neuronal ceroid lipofuscinosis. In contrast, associations for Fabry and Gaucher disease were no longer significant after adjustment; this loss of effect was primarily driven by differences in comorbidity profiles, specifically valvular disease and renal failure in Fabry and coagulopathy in Gaucher (Supplementary Table 11). These results suggest that while baseline comorbidity burden accounts for the elevated odds observed in some categories, the underlying LD phenotype remains an independent driver in others.

### Hospitalized patients with LDs and matched controls exhibit comparable survival probabilities

Survival analyses showed that patients with LDs had mortality rates similar to matched controls. Among the entire COVID Matched Cohort, one-year survival was 97.0% in controls and 95.2% in patients with LDs, an absolute difference of 1.8% (Supplementary Table 12). Using a Kaplan-Meier survival estimator, we found a significant difference in one-year survival between LD and control (log-rank p < 0.001; curve not shown), though the magnitude of separation was modest (1.8%).

Among non-hospitalized patients, one-year survival was 99.3% in controls and 98.7% in patients with LDs, also indicating a modest separation (log-rank p < 0.001; Figure 4b, Supplementary Table 12). Conversely, among hospitalized patients, one-year survival was 82.0% in controls and 82.1% in patients with LDs, a negligible difference, consistent with the difference in survival curves (log-rank p = 0.358; Figure 4a, Supplementary Table 12).

**Figure 4:**
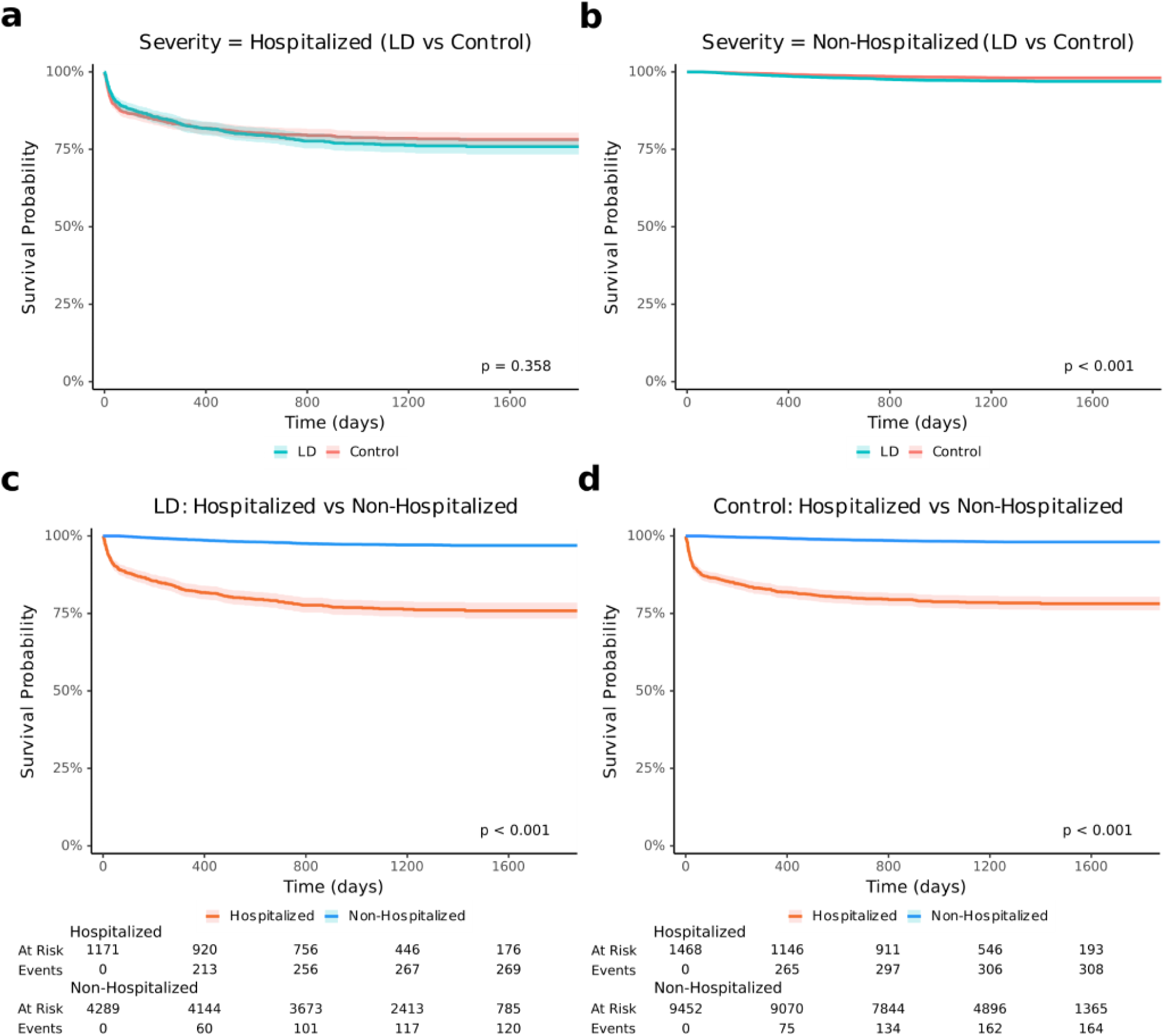
Post-COVID mortality is driven by clinical severity within cohorts rather than lysosomal disorder status between groups. Kaplan-Meier survival curves show survival probability over an 1800-day (∼5-year) follow-up period post-COVID. **a,b,** Survival trajectories for LD (teal) vs control (coral) cohorts among hospitalized patients (**a**) and non-hospitalized patients (**b**). The x-axis represents time in days post-COVID diagnosis, while the y-axis is the survival probability. **c,d,** Survival trajectories for hospitalized (orange) vs non-hospitalized (blue) individuals within the LD population (**c**) and the control (**d**) group. Risk tables underneath panels **c** and **d** indicate the number of individuals at risk and cumulative events (deaths) at 400-day intervals (0-1600 days). Shaded regions represent 95% confidence intervals. Censor marks are not shown. The p-values correspond to a two-sided log-rank test.

Within both the LD and control groups, there was a substantial difference in one-year survival between hospitalized and non-hospitalized patients, a gap exceeding 16%, also seen in the survival curves (log-rank p < 0.001; Figures 4c,d). These data indicate that while LD is associated with increased susceptibility to severe infection necessitating admission, LD status does not further modify survival trajectories once this clinical threshold is reached.

Although survival rates were similar, the timing of deaths differed markedly between the hospitalized and non-hospitalized groups. For hospitalized patients, mortality was heavily concentrated within the first 100 days following COVID-19 diagnosis, with a median time-to-death of 87 days for patients with LDs compared to 40 days for matched controls; this temporal pattern was not observed in non-hospitalized patients (Supplementary Figure 6). After this acute phase, their long-term mortality probability begins to look similar to or lower than that of patients who were never hospitalized.

### Acute-phase interventions reveal LD-dependent reductions in mortality hazards

To assess the impact of infection-modifying treatments administered during the initial stages of infection, we utilized Cox proportional hazards regression to model survival following drug and procedure administration within the first 14 days of the COVID-19 window. While overall mortality rates between LD and controls were statistically similar, the association between specific interventions and mortality differed between groups.

We noted evidence of confounding by indication, characterized by high hazard ratios (HR > 6) for several standard-of-care COVID-19 treatments and procedures in the control group, reflecting their use in clinically severe cases. However, we identified significant interaction effects indicating that these associations were LD-dependent. Specifically, the association between mortality and the administration of dexamethasone (HR^interaction^ = 0.43, 95% CI: 0.20-0.91, p = 0.027) and remdesivir (HR^interaction^ = 0.42, 95% CI: 0.19-0.95, p = 0.037) was significantly reduced in the LD cohort compared to controls (Supplementary Figure 7). Similarly, various clinical procedures, such as therapeutic infusions and injections, and aerosol or nebulizer inhalation treatments (HR^interaction^ = 0.28, 95% CI: 0.12-0.68, p = 0.005, Supplementary Figure 8), showed a reduction in mortality hazards in the LD cohort.

## Discussion

This study represents the largest investigation to date into the relationship between lysosomal disorders and severe COVID-19 outcomes, and to our knowledge, one of the largest studies of LDs overall. Using the National Clinical Cohort Collaborative COVID Enclave, we identified 16,313 patients with LDs, of whom 5,460 were PCR/antigen- or ICD-confirmed for COVID-19 and met the inclusion criteria.

First, we used multiple correspondence analysis to characterize the latent structure of comorbidity burden. This analysis revealed a distinct cardiovascular-utilization-comorbidity burden gradient along Dimension 1, driven by severe end-organ damage conditions such as congestive heart failure, complicated hypertension, and pulmonary circulation disorders. LD status and hospitalization clustered toward the high-burden end of the latent space, suggesting that LD creates a vascular-pulmonary comorbidity phenotype that exacerbates the standard COVID-19 hospitalization signature.

While the MCA defined the structure of the comorbidity burden, the prevalence differences explained how sickness changed across the severity spectrum. Patients with LDs had a significantly higher baseline comorbidity burden with a median Elixhauser score of 12, compared to 2 in matched controls. Although patients with LDs have a higher burden overall, the largest difference in mean comorbidity prevalence was in outpatients requiring an emergency department visit. The earlier observed high prevalence of cardiometabolic and pulmonary comorbidity categories shows that LDs’ baseline pattern is similar to that of much higher severity controls.

We overcame the limitations of prior small, single-center, or subtype-specific reports by using a matched cohort design and conditional logistic regression, and adjusted for comorbidities, which allowed us to evaluate associations after accounting for comorbidity burden. Comorbidity burden explains part, but not all, of the risk: hospitalization odds remained elevated overall and in several categories, indicating an independent LD effect. Notably, associations for Fabry and Gaucher disease were no longer significant; this full loss of effect was driven by valvular disease and renal failure in Fabry, and by coagulopathy in Gaucher.

The heterogeneity in adjusted risk across LD categories is consistent with different lysosomal roles in SARS-CoV-2 pathogenesis. The strong residual risk observed in neurodegenerative LDs, like neuronal ceroid lipofuscinosis and metachromatic leukodystrophy, most likely reflects mechanisms downstream of viral pathogenesis rather than altered viral entry, including dysphagia-driven aspiration pneumonia, impaired airway clearance, and reduced respiratory reserve secondary to progressive neurologic decline [15–16]. The full elimination of association with hospitalization in other categories further underscores distinct, condition-specific mechanisms. While clinical data in Gaucher disease remain largely inconclusive [7], the baseline risk in Fabry disease may be driven primarily by immunosuppressive therapies rather than by intrinsic lysosomal susceptibility [8,9]. In Fabry disease, these therapies are typically indicated for post-transplantation immunosuppression following renal failure or for immune modulation to manage high antibody titers against enzyme replacement therapy. Furthermore, the absence of independent risk in Fabry disease is consistent with mechanistic models suggesting that impaired ACE2 glycosylation and reduced Cathepsin L activity disrupt endosomal viral entry [6].

Finally, we distinguished between hospitalization and mortality. While patients with LDs were more likely to be hospitalized, one-year survival among hospitalized patients was comparable to matched controls, indicating that LD status does not independently worsen post-hospitalization survival. Rather, mortality appears driven primarily by the need for hospitalization itself.

Associations between mortality and standard-of-care treatments, dexamethasone and remdesivir, were diminished in the LD cohort, potentially reflecting a more proactive clinical management strategy, in which life-saving interventions are initiated earlier in the course of the infection. Alternatively, it remains possible that lysosomal dysfunction alters the biological response to these therapies, potentially blunting the acute inflammatory phase of the infection.

There are limitations to this study worth noting. First, longitudinal data is recorded on or after January 1st, 2018, making it difficult to fully assess preexisting comorbidities. To mitigate these risks, we included only patients with at least one encounter before the COVID-19 diagnosis.

Second, although we applied Mahalanobis distance matching and adjusted for comorbidity covariates, residual confounding remains possible. In particular, healthcare utilization, which is significantly higher among patients with LDs, may influence the ascertainment of comorbidities and the likelihood of hospitalization, independent of infection severity. Similarly, unmeasured factors such as socioeconomic status, access to specialty care, LD disorder severity across each category, and treatment history (e.g., enzyme replacement therapy or transplant status) were not fully captured and may have contributed to the observed associations.

Third, COVID-19 severity was defined using the maximum recorded severity state, which limited inference about clinical trajectories across severity strata. We also observed substantial confounding by indication in the association between mortality and standard-of-care COVID-19 interventions. This should not be taken as evidence that these treatments are harmful, but rather as an exploratory finding suggesting that LD status may modify treatment response in ways not captured in this study.

Despite these limitations, this study provides a robust, large-scale assessment of COVID-19 outcomes in patients with lysosomal disorders, offering novel insights into the independent, heterogeneous effects of lysosomal disorders on COVID-19 severity.

## Conclusions

In summary, patients with lysosomal disorders were at increased odds of COVID-19 hospitalization, driven by both elevated comorbidity burden and independent effects of the underlying disorders, with heterogeneous burden across categories. However, once hospitalized, survival outcomes were comparable to those of matched controls, indicating that the excess risk is concentrated in the transition to hospitalization. Overall, this study represents the largest and most comprehensive analysis of the effects of lysosomal disorders on COVID-19 severity outcomes.

Importantly, this study clarifies previous inconsistent and underpowered findings on the burden of severe COVID-19 in specific lysosomal disorder categories, providing clinicians with more reliable estimates to inform clinical care and treatment.

## Data Availability

The N3C Data Enclave is managed under the authority of the NIH; information can be found at https://ncats.nih.gov/n3c/resources. N3C data are protected, and can be accessed for COVID-related research with an approved (1) IRB protocol and (2) Data Use Request for study RP-582482. Enclave and data access instructions can be found at https://n3c.ncats.nih.gov/covid.

https://n3c.ncats.nih.gov/covid

https://github.com/blakebyer/ld_covid

## List of abbreviations

LD: lysosomal disorder;
N3C: National Clinical Cohort Collaborative;
EHR: electronic health record;
MPS: mucopolysaccharidosis;
SNOMED: Systematized Nomenclature of Medicine;
Mondo: Mondo Disease Ontology;
MCA: multiple correspondence analysis

## Declarations

### Ethics approval and consent to participate

This manuscript is in accordance with N3C data use and attribution policies; use of the data is approved under Johns Hopkins Office of Human Subjects Research - Institutional Review Board protocol #IRB00249128. The N3C Data Access Committee approved this study (DUR-D276495). The study used a de-identified, unconsented data set.

### Consent for publication

Not applicable.

### Availability of data and materials

Analysis code is available for review at https://github.com/blakebyer/ld_covid. Executable code is available in the N3C platform via access procedures described previously.

### Authors’ contributions

BKB: Conceptualization, Methodology, Formal Analysis, Investigation, Writing - Original Draft, Writing - Review & Editing. ZBD: Methodology, Writing - Review & Editing. BMM: Methodology, Writing - Review & Editing. JM: Writing - Review & Editing. LC: Writing - Review & Editing. MAH: Conceptualization, Resources, Writing - Review & Editing, Supervision, Funding Acquisition. STO: Conceptualization, Methodology, Writing - Review & Editing, Supervision, Project Administration.

## Acknowledgements

The authors thank Bryan Laraway for his guidance on Mondo Disease Ontology classification in OMOP. We are grateful to the Muenzer MPS Research and Treatment Center at the University of North Carolina for providing clinical context to our findings, and to Kristin Clinard and Lindsay Torrice for insightful discussions regarding the symptomatology and treatment of mucopolysaccharidoses.

