## Supplementary Material for "Differential COVID-19 Outcomes Across Lysosomal Disorders"

Blake K. Byer[1], Zachary Butzin-Dozier[2], Brenda M. McGrath[3], Joseph Muenzer[4], Lorne Clarke[5], Melissa A. Haendel[1], Shawn T. O'Neil[1*]

[1]Department of Genetics, University of North Carolina at Chapel Hill, Chapel Hill, North Carolina, United States
[2]Department of Pediatrics, Stanford University School of Medicine, Stanford, California, United States
[3]OCHIN, Portland, Oregon, United States
[4]Department of Pediatrics, University of North Carolina at Chapel Hill, Chapel Hill, North Carolina, United States
[5]Department of Medical Genetics, University of British Columbia, Vancouver, British Columbia, Canada

### Supplementary Figures

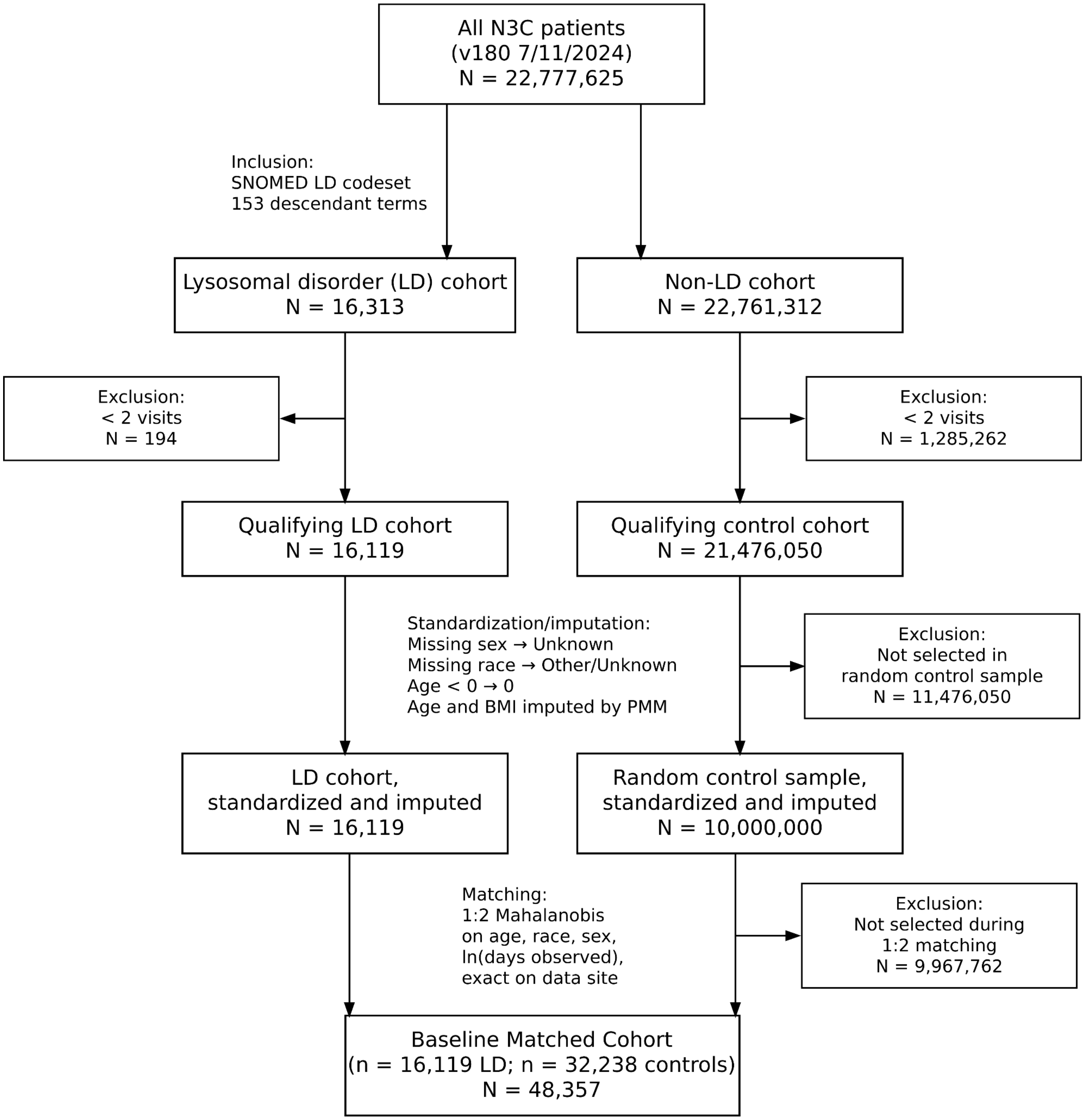

**Supplementary Figure 1: Cohort construction of the Baseline Matched Cohort.** Flowchart of inclusion and exclusion criteria applied to the N3C population. Before imputation, in LDs, age was 4.9% missing, and BMI was 11% missing; in controls, age was 4.8% missing, and BMI was 27% missing. Standardization, imputation, and 1:2 Mahalanobis matching were performed to construct a cohort comprising 16,119 patients with LDs and 32,238 controls.

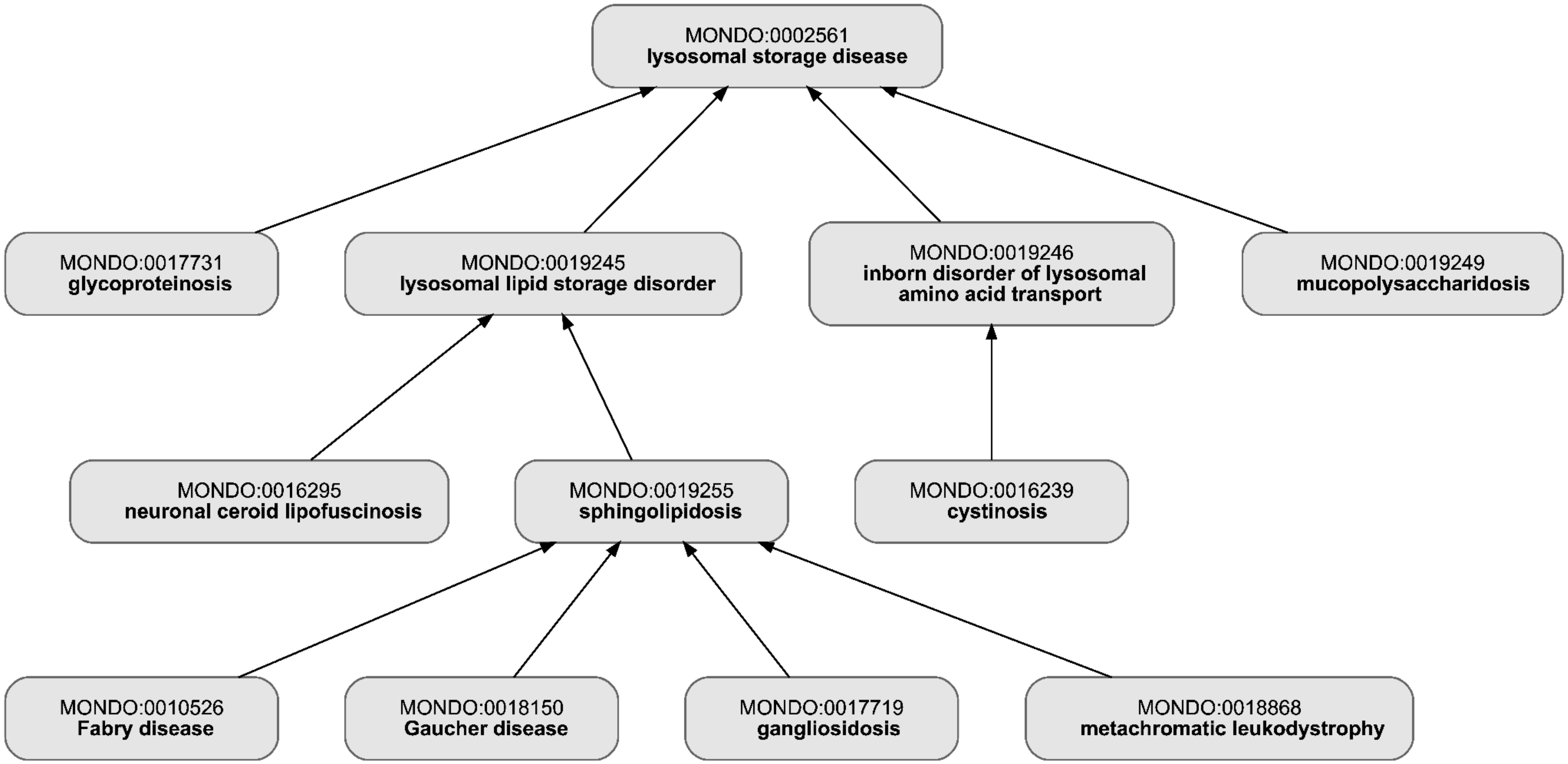

**Supplementary Figure 2: Mondo Disease Ontology lysosomal disorders hierarchy.**
Nodes represent Mondo terms (ID and label), and edges denote parent–child relationships. This hierarchy was used to define non-mutually exclusive category groupings for regression analyses.

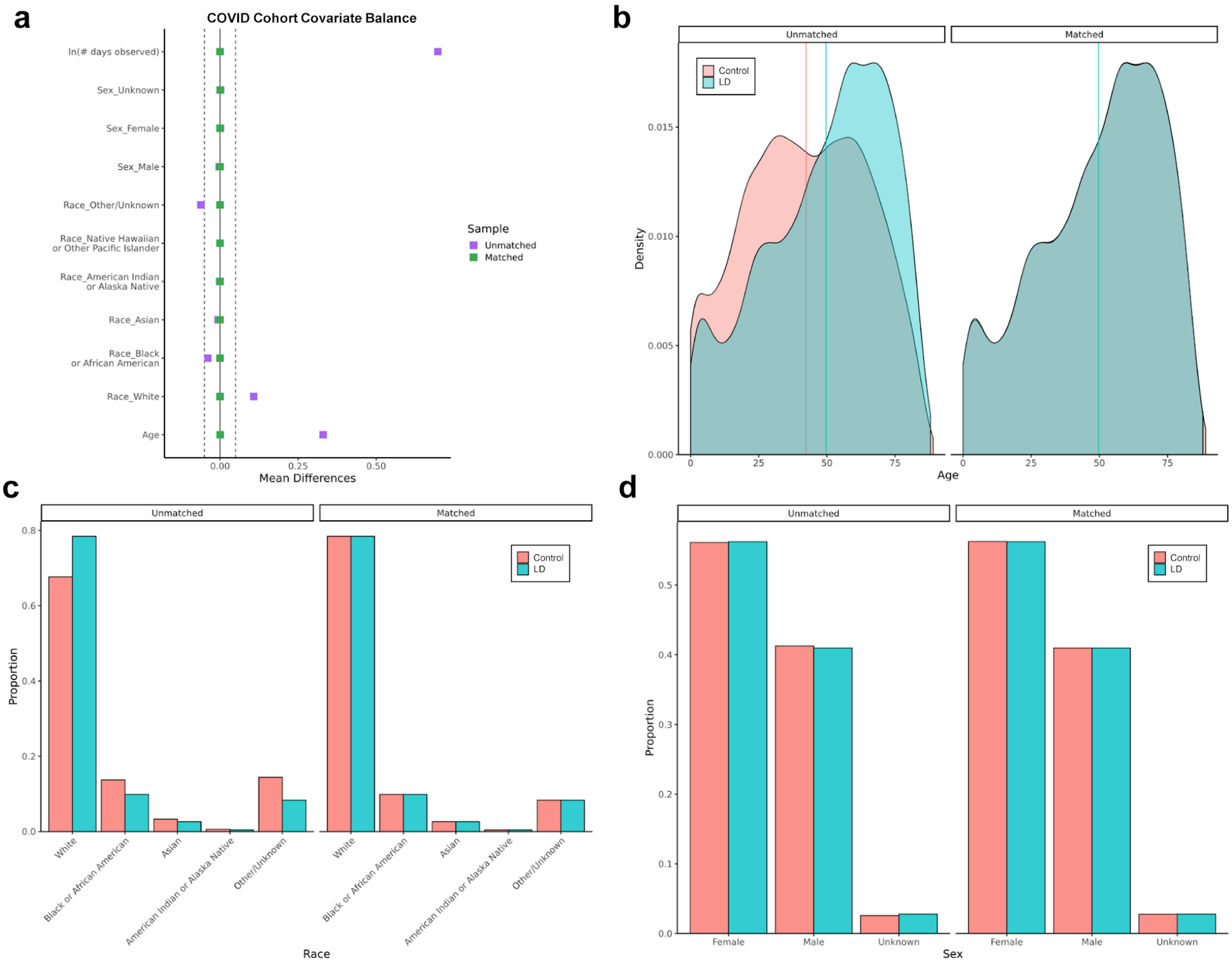
a
COVID Cohort Covariate Balance
ln(# days observed)
Sex_Unknown
Sex_Female
Sex_Male
Race_Other/Unknown
Race_Native Hawaiian or Other Pacific Islander
Race_American Indian or Alaska Native
Race_Asian
Race_Black or African American
Race_White
Age
0.00
0.25
0.50
Mean Differences
Sample
Unmatched
Matched
b
Unmatched
Matched
Control
LD
Density
0.000
0.005
0.010
0.015
0
25
50
75
Age
c
Unmatched
Matched
Control
LD
Proportion
0.0
0.2
0.4
0.6
0.8
White
Black or African American
Asian
American Indian or Alaska Native
Other/Unknown
Race
d
Unmatched
Matched
Control
LD
Proportion
0.0
0.1
0.2
0.3
0.4
0.5
Female
Male
Unknown
Sex

**Supplementary Figure 3: COVID Matched Cohort covariate balance before and after Mahalanobis distance matching**
**a,** Love plot displaying standardized mean differences (SMD) for demographic variables (age, race, sex) and a clinical utilization measure (ln[# days observed pre-COVID]) for unmatched (purple) and Mahalanobis-distance matched (green) samples. Vertical dashed lines mark the ±0.1 SMD threshold for negligible imbalance.
**b,** Age density distributions for LD (teal) and control (coral) groups, showing the reduction in age-related bias following Mahalanobis distance matching. Vertical lines indicate median age.
**c, d,** Proportional distribution of race (**c**) and sex (**d**), demonstrating the effectiveness of Mahalanobis distance matching at neutralizing demographic disparities and accounting for covariate covariance.

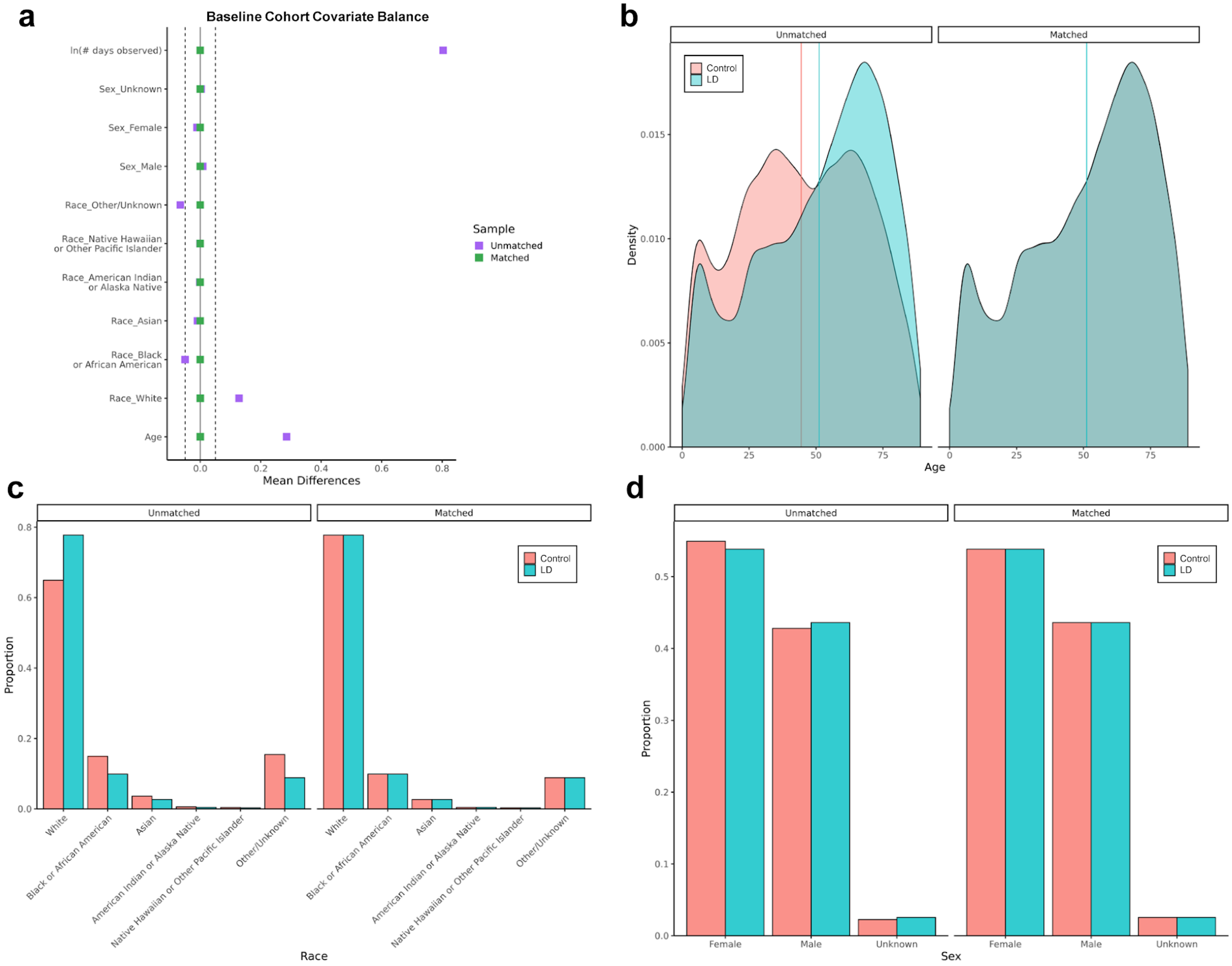
a
Baseline Cohort Covariate Balance
ln(# days observed)
Sex_Unknown
Sex_Female
Sex_Male
Race_Other/Unknown
Race_Native Hawaiian or Other Pacific Islander
Race_American Indian or Alaska Native
Race_Asian
Race_Black or African American
Race_White
Age
0.0
0.2
0.4
0.6
0.8
Mean Differences
Sample
Unmatched
Matched
b
Unmatched
Matched
Control
LD
Density
0.000
0.005
0.010
0.015
0
25
50
75
Age
c
Unmatched
Matched
Control
LD
Proportion
0.0
0.2
0.4
0.6
0.8
White
Black or African American
Asian
American Indian or Alaska Native
Native Hawaiian or Other Pacific Islander
Other/Unknown
Race
d
Unmatched
Matched
Control
LD
Proportion
0.0
0.1
0.2
0.3
0.4
0.5
Female
Male
Unknown
Sex

**Supplementary Figure 4: Baseline Matched Cohort covariate balance before and after Mahalanobis distance matching.** **a-d,** Standardized mean differences and demographic distributions for the Baseline Matched Cohort. Presentation and color schemes follow the conventions described in Supplementary Figure 3. Matching effectively neutralized disparities in age, race, and sex within the baseline population.

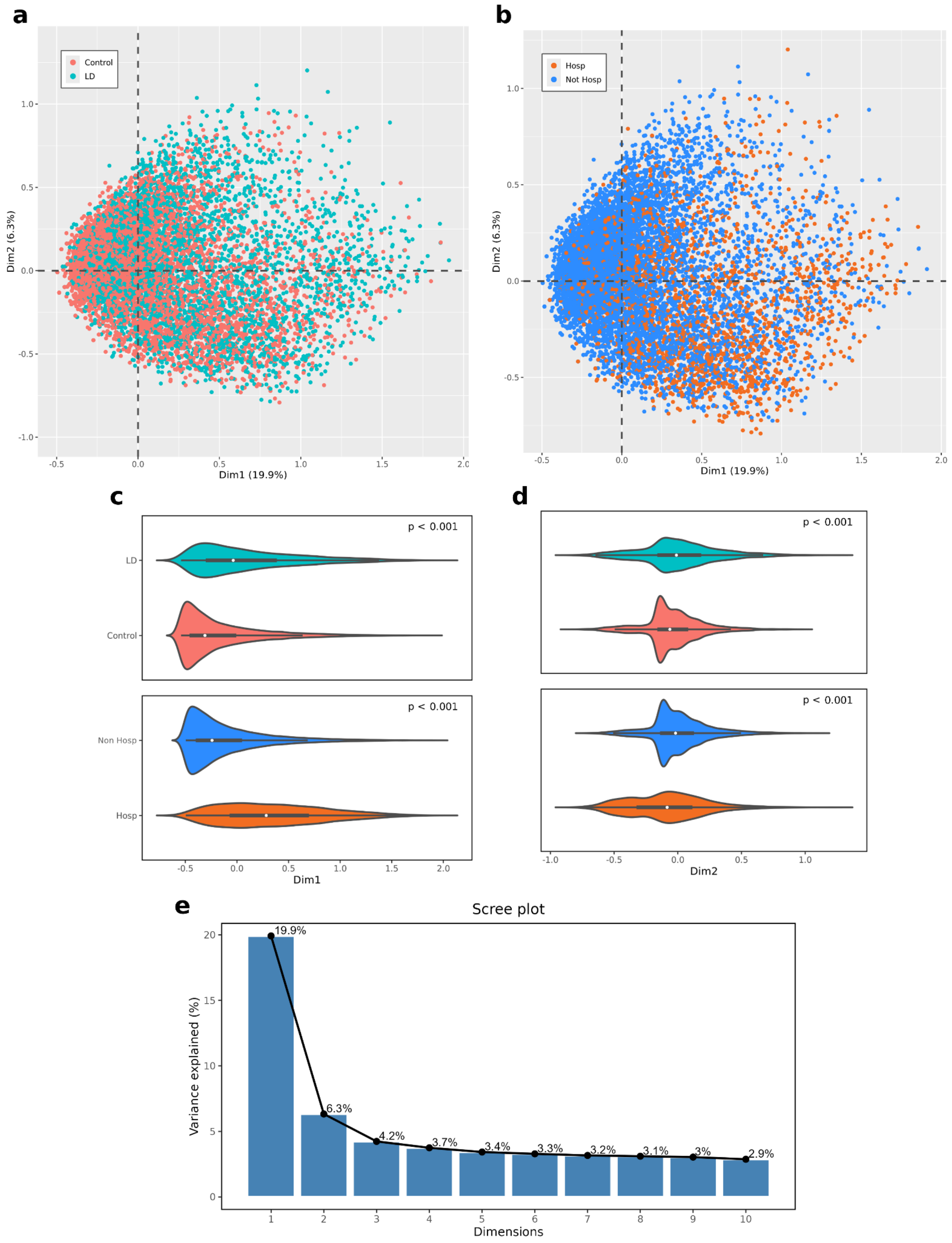

**Supplementary Figure 5: Multiple correspondence analysis of lysosomal disorder status and COVID-19 hospitalization.**

**a,** MCA score plot of the first two principal components (Dim1 and Dim2) colored by disease status (LD, teal; control, coral). **b,** Score plot colored by COVID-19 severity (hospitalized, orange; non-hospitalized, blue).

**c, d,** Marginal distributions of individuals along Dimension 1 (Dim1) (**c**) and Dimension 2 (Dim2) (**d**), stratified by lysosomal disorder status (top) and hospitalization (bottom). Violin plots illustrate the distribution of patients with median score and IQR indicated by embedded boxplots; differences between groups were assessed using a two-sided Wilcoxon rank-sum test, with LD and hospitalized patients showing significant shifts along both dimensions ($p < 0.001$).

**e,** Scree plot displaying the percentage of variance explained by the first 10 dimensions. The first dimension (Dim1) accounts for 19.9% of the total variance, while the second (Dim2) accounts for 6.3%, with a noticeable drop-off in variance captured thereafter.

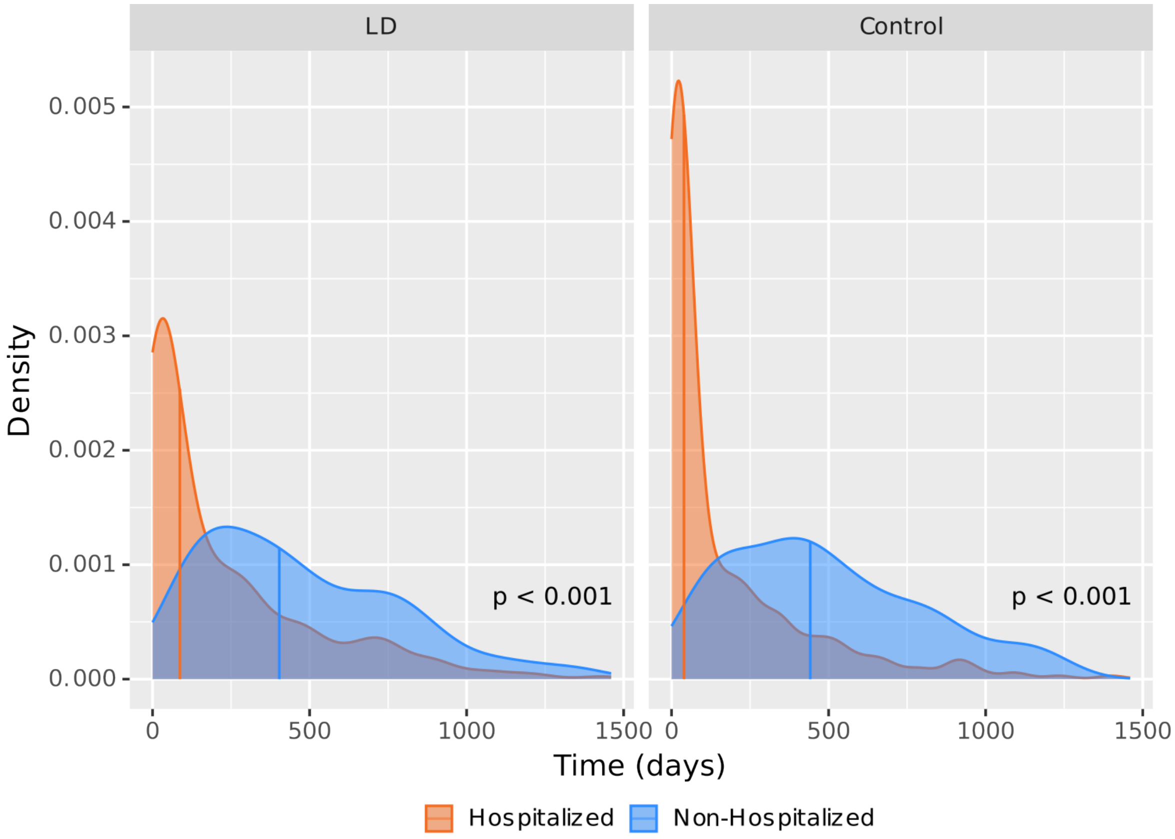

**Supplementary Figure 6: Density of Time-to-Death Post-COVID.**
Probability density distribution of time-to-death for hospitalized (orange) vs non-hospitalized (blue) across LD (left) and control (right) groups. Vertical lines indicate the median time-to-death for each group (hospitalized LD/control: 87 vs 40 days; non-hospitalized LD/control: 404 vs 442 days). P-values determined by a two-sided Wilcoxon rank-sum test comparing hospitalized and non-hospitalized patients in each panel.

| Subgroup | N | HR (95% CI) | p-value |
|---|---|---|---|
| **Albuterol** | | | |
| Drug (Control) | 821 | 3.22 (2.08, 4.98) | < 0.001 |
| Drug × LD | 653 | 0.60 (0.32, 1.13) | 0.113 |
| **Apixaban** | | | |
| Drug (Control) | 204 | 6.22 (2.73, 14.18) | < 0.001 |
| Drug × LD | 173 | 0.34 (0.11, 1.01) | 0.052 |
| **Aspirin** | | | |
| Drug (Control) | 428 | 5.47 (3.24, 9.24) | < 0.001 |
| Drug × LD | 352 | 0.32 (0.15, 0.69) | 0.004 |
| **Budesonide** | | | |
| Drug (Control) | 140 | 6.00 (2.24, 16.04) | < 0.001 |
| Drug × LD | 156 | 0.31 (0.08, 1.24) | 0.098 |
| **Carvedilol** | | | |
| Drug (Control) | 110 | 2.86 (0.92, 8.89) | 0.069 |
| Drug × LD | 112 | 1.38 (0.24, 8.07) | 0.720 |
| **Dexamethasone** | | | |
| Drug (Control) | 525 | 6.77 (3.99, 11.50) | < 0.001 |
| Drug × LD | 423 | 0.43 (0.20, 0.91) | 0.027 |
| **Famotidine** | | | |
| Drug (Control) | 382 | 3.17 (1.72, 5.84) | < 0.001 |
| Drug × LD | 284 | 0.66 (0.25, 1.69) | 0.383 |
| **Heparin** | | | |
| Drug (Control) | 271 | 6.58 (3.28, 13.20) | < 0.001 |
| Drug × LD | 257 | 0.57 (0.22, 1.48) | 0.249 |
| **Insulin** | | | |
| Drug (Control) | 430 | 6.27 (3.63, 10.81) | < 0.001 |
| Drug × LD | 404 | 0.74 (0.34, 1.59) | 0.441 |
| **Lisinopril** | | | |
| Drug (Control) | 233 | 4.89 (2.10, 11.38) | < 0.001 |
| Drug × LD | 146 | 0.47 (0.12, 1.92) | 0.294 |
| **Methylprednisolone** | | | |
| Drug (Control) | 129 | 3.08 (1.06, 8.96) | 0.039 |
| Drug × LD | 118 | 0.67 (0.16, 2.82) | 0.586 |
| **Nirmatrelvir** | | | |
| Drug (Control) | 189 | 0.48 (0.10, 2.29) | 0.360 |
| Drug × LD | 91 | 2.24 (0.17, 28.66) | 0.535 |
| **Prednisone** | | | |
| Drug (Control) | 275 | 1.36 (0.63, 2.92) | 0.432 |
| Drug × LD | 216 | 2.97 (0.89, 9.95) | 0.077 |
| **Remdesivir** | | | |
| Drug (Control) | 357 | 5.01 (2.82, 8.93) | < 0.001 |
| Drug × LD | 297 | 0.42 (0.19, 0.95) | 0.037 |
| **Systemic Hydrocortisone** | | | |
| Drug (Control) | 176 | 2.59 (0.80, 8.39) | 0.112 |
| Drug × LD | 130 | 0.34 (0.08, 1.48) | 0.151 |

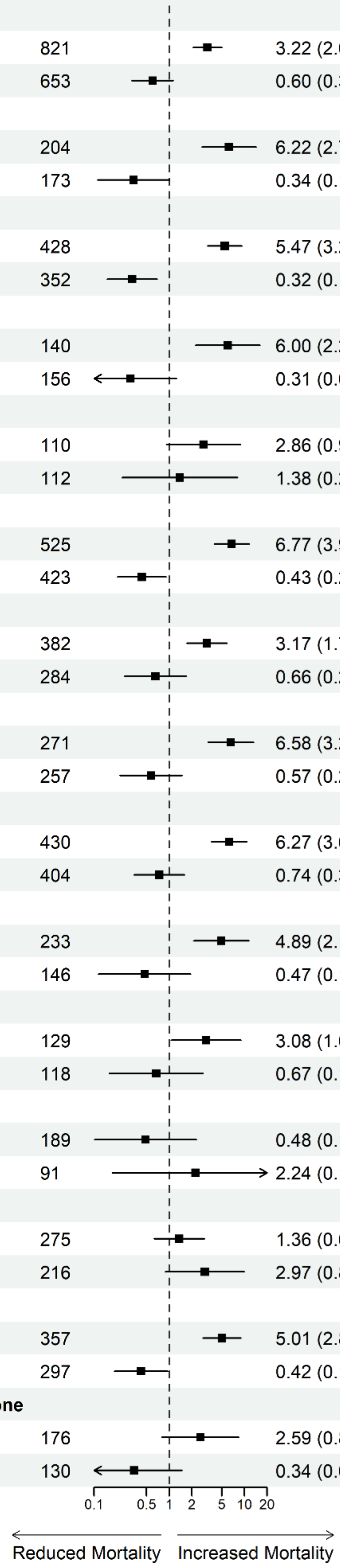

**Supplementary Figure 7: Association between acute-phase drugs and mortality with LD interaction effects.**

Forest plot showing hazard ratios (HR) and 95% confidence intervals from Cox proportional hazards models evaluating the association between administration of selected drugs and mortality. Estimates are shown for control patients (“Drug (Control)”) and for the interaction term between drug exposure and LD status (“Drug × LD”). Models were adjusted for Elixhauser comorbidity score and stratified by matched subclass. The vertical dashed line indicates HR = 1. Sample sizes (N) are displayed; for “Drug × LD,” N reflects the number of patients with LDs receiving the drug and does not necessarily correspond to the effective sample size used in model estimation. Corresponding p-values are shown.

| Subgroup | N | HR (95% CI) | p-value |
|---|---|---|---|
| **Continuous Positive Airway Pressure** | | | |
| Procedure (Control) | 106 | 20.66 (6.53, 65.35) | < 0.001 |
| Procedure × LD | 115 | 0.26 (0.05, 1.52) | 0.135 |
| **Critical Care** | | | |
| Procedure (Control) | 142 | 9.45 (4.06, 22.02) | < 0.001 |
| Procedure × LD | 159 | 0.51 (0.13, 1.98) | 0.327 |
| **Echocardiography** | | | |
| Procedure (Control) | 272 | 6.56 (3.38, 12.73) | < 0.001 |
| Procedure × LD | 249 | 0.41 (0.15, 1.09) | 0.074 |
| **Electrocardiography** | | | |
| Procedure (Control) | 728 | 4.65 (3.07, 7.06) | < 0.001 |
| Procedure × LD | 570 | 0.45 (0.24, 0.84) | 0.013 |
| **Inhalation Treatment with Aerosol or Nebulizer** | | | |
| Procedure (Control) | 261 | 10.47 (5.32, 20.61) | < 0.001 |
| Procedure × LD | 255 | 0.28 (0.12, 0.68) | 0.005 |
| **Intravenous Infusion, for Therapy or Prophylaxis** | | | |
| Procedure (Control) | 171 | 4.59 (2.00, 10.53) | < 0.001 |
| Procedure × LD | 174 | 0.27 (0.08, 0.86) | 0.027 |
| **Intravenous Infusion, Hydration** | | | |
| Procedure (Control) | 125 | 11.70 (3.37, 40.56) | < 0.001 |
| Procedure × LD | 115 | 0.07 (0.02, 0.34) | < 0.001 |
| **Other Respiratory Ventilation** | | | |
| Procedure (Control) | 56 | 9.81 (2.88, 33.38) | < 0.001 |
| Procedure × LD | 74 | 0.29 (0.05, 1.67) | 0.167 |
| **Radiologic Examination, Chest** | | | |
| Procedure (Control) | 730 | 4.99 (3.28, 7.61) | < 0.001 |
| Procedure × LD | 594 | 0.66 (0.35, 1.23) | 0.191 |
| **Specimen Collection for SARS-CoV-2** | | | |
| Procedure (Control) | 157 | 7.42 (2.35, 23.38) | < 0.001 |
| Procedure × LD | 129 | 0.24 (0.05, 1.17) | 0.077 |
| **Therapeutic, Prophylactic, or Diagnostic Injection** | | | |
| Procedure (Control) | 344 | 7.50 (3.83, 14.69) | < 0.001 |
| Procedure × LD | 300 | 0.13 (0.05, 0.36) | < 0.001 |

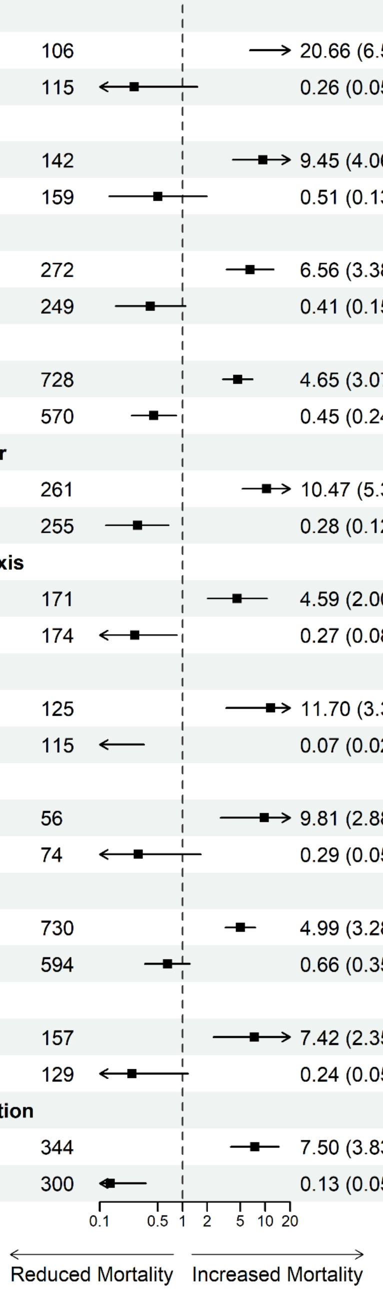

**Supplementary Figure 8: Association between acute-phase procedures and mortality with LD interaction effects.**

Forest plot showing hazard ratios (HR) and 95% confidence intervals from Cox proportional hazards models evaluating the association between administration of selected procedures and mortality. Estimates are shown for control patients ("Procedure (Control)") and for the interaction term between procedure exposure and LD status ("Procedure × LD"). Models were adjusted for Elixhauser comorbidity score and stratified by matched subclass. The vertical dashed line indicates HR = 1. Sample sizes (N) are displayed; for "Procedure × LD," N reflects the number of patients with LDs receiving the procedure and does not necessarily correspond to the effective sample size used in model estimation. Corresponding p-values are shown.

### Supplementary Tables

| MONDO Parent ID | MONDO Parent Label | SNOMED ID | SNOMED Label |
|---|---|---|---|
| MONDO:0019245 | lysosomal lipid storage disorder | SCTID:37312166 | ATPase cation transporting 13A2 related juvenile neuronal ceroid lipofuscinosis |
| MONDO:0019245 | lysosomal lipid storage disorder | SCTID:4047345 | Acute neuronopathic Gaucher's disease |
| MONDO:0019245 | lysosomal lipid storage disorder | SCTID:4029116 | Adult GM1 gangliosidosis |
| MONDO:0019245 | lysosomal lipid storage disorder | SCTID:4175778 | Adult chronic GM2 gangliosidosis |
| MONDO:0019245 | lysosomal lipid storage disorder | SCTID:4265641 | Adult neuronal ceroid lipofuscinosis |
| MONDO:0019245 | lysosomal lipid storage disorder | SCTID:4029879 | Arylsulfatase A deficiency |
| MONDO:0019245 | lysosomal lipid storage disorder | SCTID:608054 | Atypical Gaucher disease due to saposin C deficiency |
| MONDO:0019245 | lysosomal lipid storage disorder | SCTID:37111243 | Autosomal dominant myoglobinuria |
| MONDO:0019245 | lysosomal lipid storage disorder | SCTID:35622036 | Autosomal recessive cerebellar ataxia with late-onset spasticity |
| MONDO:0019245 | lysosomal lipid storage disorder | SCTID:4104697 | B variant hexosaminidase A deficiency |
| MONDO:0019245 | lysosomal lipid storage disorder | SCTID:4079872 | B variant hexosaminidase A deficiency - adult |
| MONDO:0019245 | lysosomal lipid storage disorder | SCTID:4029878 | B variant hexosaminidase A deficiency - infantile |
| MONDO:0019245 | lysosomal lipid storage disorder | SCTID:4031809 | B variant hexosaminidase A deficiency - juvenile |
| MONDO:0019245 | lysosomal lipid storage disorder | SCTID:4029115 | B1 variant hexosaminidase A deficiency |
| MONDO:0019245 | lysosomal lipid storage disorder | SCTID:374906 | Cerebral lipidosis |
| MONDO:0019245 | lysosomal lipid storage disorder | SCTID:4006310 | Chemically-induced lipidosis |
| MONDO:0019245 | lysosomal lipid storage disorder | SCTID:4210997 | Cholesterol ester storage disease |
| MONDO:0019245 | lysosomal lipid storage disorder | SCTID:4265894 | Chronic non-neuropathic Gaucher's disease |
| MONDO:0019245 | lysosomal lipid storage disorder | SCTID:36715284 | Congenital neuronal ceroid lipofuscinosis |
| MONDO:0019245 | lysosomal lipid storage disorder | SCTID:37166936 | Dystonia due to metachromatic leucodystrophy |
| MONDO:0019245 | lysosomal lipid storage disorder | SCTID:36715313 | Encephalopathy due to prosaposin deficiency |
| MONDO:0019245 | lysosomal lipid storage disorder | SCTID:4042934 | Fabry's disease |
| MONDO:0019245 | lysosomal lipid storage disorder | SCTID:4079873 | GM1 gangliosidosis |
| MONDO:0019245 | lysosomal lipid storage disorder | SCTID:4142635 | GM2 gangliosidosis |
| MONDO:0019245 | lysosomal lipid storage disorder | SCTID:4029117 | Galactocerebroside beta-galactosidase deficiency - early onset |
| MONDO:0019245 | lysosomal lipid storage disorder | SCTID:4105341 | Galactosylceramide beta-galactosidase deficiency |
| MONDO:0019245 | lysosomal lipid storage disorder | SCTID:4180066 | Gangliosidosis |
| MONDO:0019245 | lysosomal lipid storage disorder | SCTID:608071 | Gaucher disease with ophthalmoplegia and cardiovascular calcification |
| MONDO:0019245 | lysosomal lipid storage disorder | SCTID:4101305 | Gaucher's disease |
| MONDO:0019245 | lysosomal lipid storage disorder | SCTID:4294298 | Genetic disorder of lipid storage |
| MONDO:0019245 | lysosomal lipid storage disorder | SCTID:37396784 | Genetic recurrent myoglobinuria |
| MONDO:0019245 | lysosomal lipid storage disorder | SCTID:4165746 | Globoid cell leukodystrophy, late-onset |
| MONDO:0019245 | lysosomal lipid storage disorder | SCTID:4322172 | I-cell disease |
| MONDO:0019245 | lysosomal lipid storage disorder | SCTID:4031810 | Infantile GM1 gangliosidosis |
| MONDO:0019245 | lysosomal lipid storage disorder | SCTID:4266669 | Infantile GM2 gangliosidosis |
| MONDO:0019245 | lysosomal lipid storage disorder | SCTID:4241463 | Infantile neuronal ceroid lipofuscinosis |
| MONDO:0019245 | lysosomal lipid storage disorder | SCTID:4077298 | Juvenile GM1 gangliosidosis |
| MONDO:0019245 | lysosomal lipid storage disorder | SCTID:4318385 | Juvenile GM2 gangliosidosis |
| MONDO:0019245 | lysosomal lipid storage disorder | SCTID:4264152 | Juvenile neuronal ceroid lipofuscinosis |
| MONDO:0019245 | lysosomal lipid storage disorder | SCTID:4033084 | Late-infantile neuronal ceroid lipofuscinosis |
| MONDO:0019245 | lysosomal lipid storage disorder | SCTID:4027782 | Lipid storage disease |
| MONDO:0019245 | lysosomal lipid storage disorder | SCTID:4345570 | Lipid storage myopathy |
| MONDO:0019245 | lysosomal lipid storage disorder | SCTID:37399482 | Lysosomal acid lipase deficiency |
| MONDO:0019245 | lysosomal lipid storage disorder | SCTID:4262609 | Metachromatic leukodystrophy |
| MONDO:0019245 | lysosomal lipid storage disorder | SCTID:4180038 | Metachromatic leukodystrophy due to deficiency of cerebroside sulfatase activator |
| MONDO:0019245 | lysosomal lipid storage disorder | SCTID:604586 | Metachromatic leukodystrophy due to sphingolipid activator protein I deficiency |

| | | | |
|---|---|---|---|
| MONDO:0019245 | lysosomal lipid storage disorder | SCTID:4308755 | Metachromatic leukodystrophy without arylsulfatase deficiency |
| MONDO:0019245 | lysosomal lipid storage disorder | SCTID:4085741 | Metachromatic leukodystrophy, adult type |
| MONDO:0019245 | lysosomal lipid storage disorder | SCTID:4238363 | Metachromatic leukodystrophy, congenital type |
| MONDO:0019245 | lysosomal lipid storage disorder | SCTID:4185906 | Metachromatic leukodystrophy, juvenile type |
| MONDO:0019245 | lysosomal lipid storage disorder | SCTID:4166222 | Metachromatic leukodystrophy, late infantile type |
| MONDO:0019245 | lysosomal lipid storage disorder | SCTID:4183021 | Multiple sulfatase deficiency |
| MONDO:0019245 | lysosomal lipid storage disorder | SCTID:4170931 | Neuronal ceroid lipofuscinosis |
| MONDO:0019245 | lysosomal lipid storage disorder | SCTID:45771339 | Neuronal ceroid lipofuscinosis 8 |
| MONDO:0019245 | lysosomal lipid storage disorder | SCTID:44783251 | Neutral lipid storage disease with myopathy |
| MONDO:0019245 | lysosomal lipid storage disorder | SCTID:4181427 | Niemann-Pick disease, type A |
| MONDO:0019245 | lysosomal lipid storage disorder | SCTID:4214475 | Niemann-Pick disease, type B |
| MONDO:0019245 | lysosomal lipid storage disorder | SCTID:4283682 | Niemann-Pick disease, type C |
| MONDO:0019245 | lysosomal lipid storage disorder | SCTID:4314140 | Niemann-Pick disease, type C, acute form |
| MONDO:0019245 | lysosomal lipid storage disorder | SCTID:4219251 | Niemann-Pick disease, type C, chronic form |
| MONDO:0019245 | lysosomal lipid storage disorder | SCTID:4283951 | Niemann-Pick disease, type C, subacute form |
| MONDO:0019245 | lysosomal lipid storage disorder | SCTID:4078073 | Niemann-Pick disease, type D |
| MONDO:0019245 | lysosomal lipid storage disorder | SCTID:3663236 | Perinatal lethal Gaucher disease |
| MONDO:0019245 | lysosomal lipid storage disorder | SCTID:37168357 | Primary triglyceride deposit cardiomyovasculopathy |
| MONDO:0019245 | lysosomal lipid storage disorder | SCTID:37204409 | Progressive myoclonic epilepsy type 3 |
| MONDO:0019245 | lysosomal lipid storage disorder | SCTID:4119434 | Pulmonary lipid storage disease |
| MONDO:0019245 | lysosomal lipid storage disorder | SCTID:439689 | Retinal dystrophy in cerebroretinal lipidosis |
| MONDO:0019245 | lysosomal lipid storage disorder | SCTID:443895 | Retinal dystrophy in systemic lipidosis |
| MONDO:0019245 | lysosomal lipid storage disorder | SCTID:4314414 | Salla disease |
| MONDO:0019245 | lysosomal lipid storage disorder | SCTID:4031489 | Sandhoff disease |
| MONDO:0019245 | lysosomal lipid storage disorder | SCTID:4286511 | Sphingolipid activator protein 1 deficiency |
| MONDO:0019245 | lysosomal lipid storage disorder | SCTID:4079874 | Sphingolipidosis |
| MONDO:0019245 | lysosomal lipid storage disorder | SCTID:4242275 | Sphingomyelin/cholesterol lipidosis |
| MONDO:0019245 | lysosomal lipid storage disorder | SCTID:4242740 | Subacute neuronopathic Gaucher's disease |
| MONDO:0019245 | lysosomal lipid storage disorder | SCTID:4009170 | Tay-Sachs disease |
| MONDO:0019245 | lysosomal lipid storage disorder | SCTID:4324845 | Tay-Sachs disease, variant AB |
| MONDO:0019245 | lysosomal lipid storage disorder | SCTID:4029114 | Total hexosaminidase deficiency - adult |
| MONDO:0019245 | lysosomal lipid storage disorder | SCTID:4029113 | Total hexosaminidase deficiency - infantile |
| MONDO:0019245 | lysosomal lipid storage disorder | SCTID:4029752 | Total hexosaminidase deficiency - juvenile |
| MONDO:0019245 | lysosomal lipid storage disorder | SCTID:4024103 | Triglyceride storage disease with ichthyosis |
| MONDO:0019245 | lysosomal lipid storage disorder | SCTID:4221254 | Wolman's disease |
| MONDO:0019245 | lysosomal lipid storage disorder | SCTID:4029257 | Xanthomatosis, familial |
| MONDO:0019255 | sphingolipidosis | SCTID:4047345 | Acute neuronopathic Gaucher's disease |
| MONDO:0019255 | sphingolipidosis | SCTID:4029116 | Adult GM1 gangliosidosis |
| MONDO:0019255 | sphingolipidosis | SCTID:4175778 | Adult chronic GM2 gangliosidosis |
| MONDO:0019255 | sphingolipidosis | SCTID:4029879 | Arylsulfatase A deficiency |
| MONDO:0019255 | sphingolipidosis | SCTID:608054 | Atypical Gaucher disease due to saposin C deficiency |
| MONDO:0019255 | sphingolipidosis | SCTID:35622036 | Autosomal recessive cerebellar ataxia with late-onset spasticity |
| MONDO:0019255 | sphingolipidosis | SCTID:4104697 | B variant hexosaminidase A deficiency |
| MONDO:0019255 | sphingolipidosis | SCTID:4079872 | B variant hexosaminidase A deficiency - adult |
| MONDO:0019255 | sphingolipidosis | SCTID:4029878 | B variant hexosaminidase A deficiency - infantile |
| MONDO:0019255 | sphingolipidosis | SCTID:4031809 | B variant hexosaminidase A deficiency - juvenile |
| MONDO:0019255 | sphingolipidosis | SCTID:4029115 | B1 variant hexosaminidase A deficiency |
| MONDO:0019255 | sphingolipidosis | SCTID:4265894 | Chronic non-neuropathic Gaucher's disease |
| MONDO:0019255 | sphingolipidosis | SCTID:37166936 | Dystonia due to metachromatic leucodystrophy |
| MONDO:0019255 | sphingolipidosis | SCTID:36715313 | Encephalopathy due to prosaposin deficiency |
| MONDO:0019255 | sphingolipidosis | SCTID:4042934 | Fabry's disease |
| MONDO:0019255 | sphingolipidosis | SCTID:4196512 | Farber's lipogranulomatosis |
| MONDO:0019255 | sphingolipidosis | SCTID:4079873 | GM1 gangliosidosis |
| MONDO:0019255 | sphingolipidosis | SCTID:4142635 | GM2 gangliosidosis |
| MONDO:0019255 | sphingolipidosis | SCTID:4029117 | Galactocerebroside beta-galactosidase deficiency - early onset |

| | | | |
|---|---|---|---|
| MONDO:0019255 | sphingolipidosis | SCTID:4105341 | Galactosylceramide beta-galactosidase deficiency |
| MONDO:0019255 | sphingolipidosis | SCTID:4180066 | Gangliosidosis |
| MONDO:0019255 | sphingolipidosis | SCTID:608071 | Gaucher disease with ophthalmoplegia and cardiovascular calcification |
| MONDO:0019255 | sphingolipidosis | SCTID:4101305 | Gaucher's disease |
| MONDO:0019255 | sphingolipidosis | SCTID:4165746 | Globoid cell leukodystrophy, late-onset |
| MONDO:0019255 | sphingolipidosis | SCTID:4031810 | Infantile GM1 gangliosidosis |
| MONDO:0019255 | sphingolipidosis | SCTID:4266669 | Infantile GM2 gangliosidosis |
| MONDO:0019255 | sphingolipidosis | SCTID:4077298 | Juvenile GM1 gangliosidosis |
| MONDO:0019255 | sphingolipidosis | SCTID:4318385 | Juvenile GM2 gangliosidosis |
| MONDO:0019255 | sphingolipidosis | SCTID:4262609 | Metachromatic leukodystrophy |
| MONDO:0019255 | sphingolipidosis | SCTID:4180038 | Metachromatic leukodystrophy due to deficiency of cerebroside sulfatase activator |
| MONDO:0019255 | sphingolipidosis | SCTID:604586 | Metachromatic leukodystrophy due to sphingolipid activator protein I deficiency |
| MONDO:0019255 | sphingolipidosis | SCTID:4308755 | Metachromatic leukodystrophy without arylsulfatase deficiency |
| MONDO:0019255 | sphingolipidosis | SCTID:4085741 | Metachromatic leukodystrophy, adult type |
| MONDO:0019255 | sphingolipidosis | SCTID:4238363 | Metachromatic leukodystrophy, congenital type |
| MONDO:0019255 | sphingolipidosis | SCTID:4185906 | Metachromatic leukodystrophy, juvenile type |
| MONDO:0019255 | sphingolipidosis | SCTID:4166222 | Metachromatic leukodystrophy, late infantile type |
| MONDO:0019255 | sphingolipidosis | SCTID:4183021 | Multiple sulfatase deficiency |
| MONDO:0019255 | sphingolipidosis | SCTID:4181427 | Niemann-Pick disease, type A |
| MONDO:0019255 | sphingolipidosis | SCTID:4214475 | Niemann-Pick disease, type B |
| MONDO:0019255 | sphingolipidosis | SCTID:4283682 | Niemann-Pick disease, type C |
| MONDO:0019255 | sphingolipidosis | SCTID:4314140 | Niemann-Pick disease, type C, acute form |
| MONDO:0019255 | sphingolipidosis | SCTID:4219251 | Niemann-Pick disease, type C, chronic form |
| MONDO:0019255 | sphingolipidosis | SCTID:4283951 | Niemann-Pick disease, type C, subacute form |
| MONDO:0019255 | sphingolipidosis | SCTID:4078073 | Niemann-Pick disease, type D |
| MONDO:0019255 | sphingolipidosis | SCTID:3663236 | Perinatal lethal Gaucher disease |
| MONDO:0019255 | sphingolipidosis | SCTID:4031489 | Sandhoff disease |
| MONDO:0019255 | sphingolipidosis | SCTID:4286511 | Sphingolipid activator protein 1 deficiency |
| MONDO:0019255 | sphingolipidosis | SCTID:4079874 | Sphingolipidosis |
| MONDO:0019255 | sphingolipidosis | SCTID:4242275 | Sphingomyelin/cholesterol lipidosis |
| MONDO:0019255 | sphingolipidosis | SCTID:4242740 | Subacute neuronopathic Gaucher's disease |
| MONDO:0019255 | sphingolipidosis | SCTID:4009170 | Tay-Sachs disease |
| MONDO:0019255 | sphingolipidosis | SCTID:4324845 | Tay-Sachs disease, variant AB |
| MONDO:0019255 | sphingolipidosis | SCTID:4029114 | Total hexosaminidase deficiency - adult |
| MONDO:0019255 | sphingolipidosis | SCTID:4029113 | Total hexosaminidase deficiency - infantile |
| MONDO:0019255 | sphingolipidosis | SCTID:4029752 | Total hexosaminidase deficiency - juvenile |
| MONDO:0010526 | Fabry disease | SCTID:4042934 | Fabry's disease |
| MONDO:0019249 | mucopolysaccharidosis | SCTID:4199300 | Deficiency of N-acetylgalactosamine-4-sulfatase |
| MONDO:0019249 | mucopolysaccharidosis | SCTID:4208985 | Hunter's syndrome, mild form |
| MONDO:0019249 | mucopolysaccharidosis | SCTID:4247774 | Hunter's syndrome, severe form |
| MONDO:0019249 | mucopolysaccharidosis | SCTID:4287262 | Maroteaux-Lamy syndrome |
| MONDO:0019249 | mucopolysaccharidosis | SCTID:4094220 | Maroteaux-Lamy syndrome, intermediate form |
| MONDO:0019249 | mucopolysaccharidosis | SCTID:4285890 | Maroteaux-Lamy syndrome, mild form |
| MONDO:0019249 | mucopolysaccharidosis | SCTID:4238503 | Maroteaux-Lamy syndrome, severe form |
| MONDO:0019249 | mucopolysaccharidosis | SCTID:4292105 | Morquio syndrome |
| MONDO:0019249 | mucopolysaccharidosis | SCTID:433446 | Mucopolysaccharidosis |
| MONDO:0019249 | mucopolysaccharidosis | SCTID:4294142 | Mucopolysaccharidosis, MPS-I |
| MONDO:0019249 | mucopolysaccharidosis | SCTID:4276347 | Mucopolysaccharidosis, MPS-I-H |
| MONDO:0019249 | mucopolysaccharidosis | SCTID:4097554 | Mucopolysaccharidosis, MPS-I-H/S |
| MONDO:0019249 | mucopolysaccharidosis | SCTID:4218929 | Mucopolysaccharidosis, MPS-I-S |
| MONDO:0019249 | mucopolysaccharidosis | SCTID:4323827 | Mucopolysaccharidosis, MPS-II |
| MONDO:0019249 | mucopolysaccharidosis | SCTID:4215209 | Mucopolysaccharidosis, MPS-III-A |
| MONDO:0019249 | mucopolysaccharidosis | SCTID:4243053 | Mucopolysaccharidosis, MPS-III-B |
| MONDO:0019249 | mucopolysaccharidosis | SCTID:4326882 | Mucopolysaccharidosis, MPS-III-C |

| | | | |
|---|---|---|---|
| MONDO:0019249 | mucopolysaccharidosis | SCTID:4049596 | Mucopolysaccharidosis, MPS-III-D |
| MONDO:0019249 | mucopolysaccharidosis | SCTID:4219277 | Mucopolysaccharidosis, MPS-IV-A |
| MONDO:0019249 | mucopolysaccharidosis | SCTID:4031815 | Mucopolysaccharidosis, MPS-IV-B |
| MONDO:0019249 | mucopolysaccharidosis | SCTID:4184782 | Mucopolysaccharidosis, MPS-VII |
| MONDO:0019249 | mucopolysaccharidosis | SCTID:37162245 | Mucopolysaccharidosis-like plus disease |
| MONDO:0019249 | mucopolysaccharidosis | SCTID:4227905 | Sanfilippo syndrome |
| MONDO:0018868 | metachromatic leukodystrophy | SCTID:4029879 | Arylsulfatase A deficiency |
| MONDO:0018868 | metachromatic leukodystrophy | SCTID:4280234 | Deficiency of cerebroside-sulfatase |
| MONDO:0018868 | metachromatic leukodystrophy | SCTID:37166936 | Dystonia due to metachromatic leucodystrophy |
| MONDO:0018868 | metachromatic leukodystrophy | SCTID:4262609 | Metachromatic leukodystrophy |
| MONDO:0018868 | metachromatic leukodystrophy | SCTID:4180038 | Metachromatic leukodystrophy due to deficiency of cerebroside sulfatase activator |
| MONDO:0018868 | metachromatic leukodystrophy | SCTID:604586 | Metachromatic leukodystrophy due to sphingolipid activator protein I deficiency |
| MONDO:0018868 | metachromatic leukodystrophy | SCTID:4308755 | Metachromatic leukodystrophy without arylsulfatase deficiency |
| MONDO:0018868 | metachromatic leukodystrophy | SCTID:4085741 | Metachromatic leukodystrophy, adult type |
| MONDO:0018868 | metachromatic leukodystrophy | SCTID:4238363 | Metachromatic leukodystrophy, congenital type |
| MONDO:0018868 | metachromatic leukodystrophy | SCTID:4185906 | Metachromatic leukodystrophy, juvenile type |
| MONDO:0018868 | metachromatic leukodystrophy | SCTID:4166222 | Metachromatic leukodystrophy, late infantile type |
| MONDO:0018868 | metachromatic leukodystrophy | SCTID:4286511 | Sphingolipid activator protein 1 deficiency |
| MONDO:0018150 | Gaucher disease | SCTID:4047345 | Acute neuronopathic Gaucher's disease |
| MONDO:0018150 | Gaucher disease | SCTID:608054 | Atypical Gaucher disease due to saposin C deficiency |
| MONDO:0018150 | Gaucher disease | SCTID:4265894 | Chronic non-neuropathic Gaucher's disease |
| MONDO:0018150 | Gaucher disease | SCTID:608071 | Gaucher disease with ophthalmoplegia and cardiovascular calcification |
| MONDO:0018150 | Gaucher disease | SCTID:4101305 | Gaucher's disease |
| MONDO:0018150 | Gaucher disease | SCTID:3663236 | Perinatal lethal Gaucher disease |
| MONDO:0018150 | Gaucher disease | SCTID:4242740 | Subacute neuronopathic Gaucher's disease |
| MONDO:0016295 | neuronal ceroid lipofuscinosis | SCTID:37312166 | ATPase cation transporting 13A2 related juvenile neuronal ceroid lipofuscinosis |
| MONDO:0016295 | neuronal ceroid lipofuscinosis | SCTID:4265641 | Adult neuronal ceroid lipofuscinosis |
| MONDO:0016295 | neuronal ceroid lipofuscinosis | SCTID:36715284 | Congenital neuronal ceroid lipofuscinosis |
| MONDO:0016295 | neuronal ceroid lipofuscinosis | SCTID:4241463 | Infantile neuronal ceroid lipofuscinosis |
| MONDO:0016295 | neuronal ceroid lipofuscinosis | SCTID:4264152 | Juvenile neuronal ceroid lipofuscinosis |
| MONDO:0016295 | neuronal ceroid lipofuscinosis | SCTID:4033084 | Late-infantile neuronal ceroid lipofuscinosis |
| MONDO:0016295 | neuronal ceroid lipofuscinosis | SCTID:4170931 | Neuronal ceroid lipofuscinosis |
| MONDO:0016295 | neuronal ceroid lipofuscinosis | SCTID:45771339 | Neuronal ceroid lipofuscinosis 8 |
| MONDO:0016295 | neuronal ceroid lipofuscinosis | SCTID:37204409 | Progressive myoclonic epilepsy type 3 |
| MONDO:0017719 | gangliosidosis | SCTID:4029116 | Adult GM1 gangliosidosis |
| MONDO:0017719 | gangliosidosis | SCTID:4175778 | Adult chronic GM2 gangliosidosis |
| MONDO:0017719 | gangliosidosis | SCTID:4104697 | B variant hexosaminidase A deficiency |
| MONDO:0017719 | gangliosidosis | SCTID:4079872 | B variant hexosaminidase A deficiency - adult |
| MONDO:0017719 | gangliosidosis | SCTID:4029878 | B variant hexosaminidase A deficiency - infantile |
| MONDO:0017719 | gangliosidosis | SCTID:4031809 | B variant hexosaminidase A deficiency - juvenile |
| MONDO:0017719 | gangliosidosis | SCTID:4029115 | B1 variant hexosaminidase A deficiency |
| MONDO:0017719 | gangliosidosis | SCTID:4079873 | GM1 gangliosidosis |
| MONDO:0017719 | gangliosidosis | SCTID:4142635 | GM2 gangliosidosis |
| MONDO:0017719 | gangliosidosis | SCTID:4180066 | Gangliosidosis |
| MONDO:0017719 | gangliosidosis | SCTID:4031810 | Infantile GM1 gangliosidosis |
| MONDO:0017719 | gangliosidosis | SCTID:4266669 | Infantile GM2 gangliosidosis |
| MONDO:0017719 | gangliosidosis | SCTID:4077298 | Juvenile GM1 gangliosidosis |
| MONDO:0017719 | gangliosidosis | SCTID:4318385 | Juvenile GM2 gangliosidosis |
| MONDO:0017719 | gangliosidosis | SCTID:4031489 | Sandhoff disease |
| MONDO:0017719 | gangliosidosis | SCTID:4009170 | Tay-Sachs disease |
| MONDO:0017719 | gangliosidosis | SCTID:4324845 | Tay-Sachs disease, variant AB |
| MONDO:0017719 | gangliosidosis | SCTID:4029114 | Total hexosaminidase deficiency - adult |
| MONDO:0017719 | gangliosidosis | SCTID:4029113 | Total hexosaminidase deficiency - infantile |

| | | | |
|---|---|---|---|
| MONDO:0017719 | gangliosidosis | SCTID:4029752 | Total hexosaminidase deficiency - juvenile |
| MONDO:0019246 | inborn disorder of lysosomal amino acid transport | SCTID:44809321 | Adult cystinosis |
| MONDO:0019246 | inborn disorder of lysosomal amino acid transport | SCTID:4094164 | Benign adult cystinosis |
| MONDO:0019246 | inborn disorder of lysosomal amino acid transport | SCTID:4126105 | Congenital Fanconi syndrome |
| MONDO:0019246 | inborn disorder of lysosomal amino acid transport | SCTID:4100827 | Cystinosis |
| MONDO:0019246 | inborn disorder of lysosomal amino acid transport | SCTID:4079879 | Disorder of sialic acid metabolism |
| MONDO:0019246 | inborn disorder of lysosomal amino acid transport | SCTID:4268774 | Infantile nephropathic cystinosis |
| MONDO:0019246 | inborn disorder of lysosomal amino acid transport | SCTID:4027350 | Juvenile nephropathic cystinosis |
| MONDO:0019246 | inborn disorder of lysosomal amino acid transport | SCTID:4314414 | Salla disease |
| MONDO:0019246 | inborn disorder of lysosomal amino acid transport | SCTID:4146367 | Sialic acid storage disease, severe infantile type |
| MONDO:0019246 | inborn disorder of lysosomal amino acid transport | SCTID:4150750 | Sialic storage disease |
| MONDO:0019246 | inborn disorder of lysosomal amino acid transport | SCTID:4031943 | Sialuria |
| MONDO:0016239 | cystinosis | SCTID:44809321 | Adult cystinosis |
| MONDO:0016239 | cystinosis | SCTID:4094164 | Benign adult cystinosis |
| MONDO:0016239 | cystinosis | SCTID:4126105 | Congenital Fanconi syndrome |
| MONDO:0016239 | cystinosis | SCTID:4100827 | Cystinosis |
| MONDO:0016239 | cystinosis | SCTID:4268774 | Infantile nephropathic cystinosis |
| MONDO:0016239 | cystinosis | SCTID:4027350 | Juvenile nephropathic cystinosis |
| MONDO:0017731 | glycoproteinosis | SCTID:4163879 | Adult fucosidosis |
| MONDO:0017731 | glycoproteinosis | SCTID:4029122 | Alpha-N-acetylgalactosaminidase deficiency |
| MONDO:0017731 | glycoproteinosis | SCTID:604334 | Alpha-N-acetylgalactosaminidase deficiency type 1 |
| MONDO:0017731 | glycoproteinosis | SCTID:619007 | Alpha-N-acetylgalactosaminidase deficiency type 2 |
| MONDO:0017731 | glycoproteinosis | SCTID:619008 | Alpha-N-acetylgalactosaminidase deficiency type 3 |
| MONDO:0017731 | glycoproteinosis | SCTID:4207521 | Aspartylglucosaminuria |
| MONDO:0017731 | glycoproteinosis | SCTID:4031942 | Beta-D-mannosidosis |
| MONDO:0017731 | glycoproteinosis | SCTID:4262271 | Combined deficiency of sialidase AND beta galactosidase |
| MONDO:0017731 | glycoproteinosis | SCTID:4181428 | Dysmorphic sialidosis |
| MONDO:0017731 | glycoproteinosis | SCTID:4217611 | Dysmorphic sialidosis with renal involvement |
| MONDO:0017731 | glycoproteinosis | SCTID:4100490 | Dysmorphic sialidosis, congenital form |
| MONDO:0017731 | glycoproteinosis | SCTID:4202220 | Dysmorphic sialidosis, infantile form |
| MONDO:0017731 | glycoproteinosis | SCTID:4008557 | Dysmorphic sialidosis, juvenile form |
| MONDO:0017731 | glycoproteinosis | SCTID:4277436 | Fucosidosis |
| MONDO:0017731 | glycoproteinosis | SCTID:4299945 | Glycoprotein storage disorder |
| MONDO:0017731 | glycoproteinosis | SCTID:4322172 | I-cell disease |
| MONDO:0017731 | glycoproteinosis | SCTID:4246880 | Infantile fucosidosis |
| MONDO:0017731 | glycoproteinosis | SCTID:4161004 | Juvenile fucosidosis |
| MONDO:0017731 | glycoproteinosis | SCTID:4277255 | Mannosidosis |
| MONDO:0017731 | glycoproteinosis | SCTID:4266511 | Mannosidosis, type I |
| MONDO:0017731 | glycoproteinosis | SCTID:4241350 | Mannosidosis, type II |
| MONDO:0017731 | glycoproteinosis | SCTID:4323268 | Mucolipidosis |
| MONDO:0017731 | glycoproteinosis | SCTID:37110837 | Mucolipidosis type IV |
| MONDO:0017731 | glycoproteinosis | SCTID:607719 | Oligosaccharidosis |
| MONDO:0017731 | glycoproteinosis | SCTID:4278525 | Pseudo-Hurler polydystrophy |
| MONDO:0017731 | glycoproteinosis | SCTID:4307744 | Sialidosis |
| MONDO:0017731 | glycoproteinosis | SCTID:37109814 | Sialidosis type 1 |
| unassigned | unassigned | SCTID:4212254 | Acid phosphatase deficiency |

| | | | |
|---|---|---|---|
| unassigned | unassigned | SCTID:4185398 | Arylsulfatase deficiency without MLD (metachromatic leukodystrophy) |
| unassigned | unassigned | SCTID:4167868 | Danon disease |
| unassigned | unassigned | SCTID:4053270 | Disorder of lysosomal enzyme |
| unassigned | unassigned | SCTID:4109172 | Dysostosis multiplex |
| unassigned | unassigned | SCTID:4150135 | Dysostosis multiplex group |
| unassigned | unassigned | SCTID:4151377 | Intestinal lipofuscinosis |
| unassigned | unassigned | SCTID:4030564 | Lipofuscinosis |
| unassigned | unassigned | SCTID:37155637 | Lysosomal storage disease |
| unassigned | unassigned | SCTID:4302604 | Pancreatic triacylglycerol lipase deficiency |

**Supplementary Table 1: Mapping of SNOMED diagnoses to MONDO parent categories.** SNOMED-coded lysosomal disorder diagnoses were mapped to higher-level MONDO parent categories based on hierarchical relationships in the Mondo Disease Ontology; assignments represent parent-child relationships and not necessarily exact synonyms or cross-references.

| Mondo ID | Mondo Label | Patient Count |
|---|---|---|
| MONDO:0019245 | lysosomal lipid storage disorder | 14654 |
| MONDO:0019255 | sphingolipidosis | 7774 |
| MONDO:0010526 | Fabry disease | 1117 |
| MONDO:0019249 | mucopolysaccharidosis | 1096 |
| MONDO:0018868 | metachromatic leukodystrophy | 467 |
| MONDO:0018150 | Gaucher disease | 455 |
| MONDO:0016295 | neuronal ceroid lipofuscinosis | 303 |
| MONDO:0017719 | gangliosidosis | 298 |
| MONDO:0019246 | inborn disorder of lysosomal amino acid transport | 240 |
| MONDO:0016239 | cystinosis | 238 |
| MONDO:0017731 | glycoproteinosis | 30 |

**Supplementary Table 2: Lysosomal Disorder Rolled-Up Category Counts in the Baseline Matched Cohort.**

| Mondo ID | Mondo Label | Patient Count |
|---|---|---|
| MONDO:0019245 | lysosomal lipid storage disorder | 5046 |
| MONDO:0019255 | sphingolipidosis | 2624 |
| MONDO:0010526 | Fabry disease | 368 |
| MONDO:0019249 | mucopolysaccharidosis | 321 |
| MONDO:0018868 | metachromatic leukodystrophy | 160 |
| MONDO:0018150 | Gaucher disease | 145 |
| MONDO:0017719 | gangliosidosis | 105 |
| MONDO:0016295 | neuronal ceroid lipofuscinosis | 99 |
| MONDO:0019246 | inborn disorder of lysosomal amino acid transport | 76 |
| MONDO:0016239 | cystinosis | 76 |
| MONDO:0017731 | glycoproteinosis | <20 |

**Supplementary Table 3: Lysosomal Disorder Rolled-Up Category Counts in the COVID Matched Cohort.**

| Codeset ID | Concept Set Name |
|---|---|
| 270102919 | [Pitt N3C] Heparin Sodium (UFH) |
| 932266800 | systemic hydrocortisone35 |
| 231875380 | famotidine-105 |
| 425433230 | [DM]Insulin |
| 1000082442 | Nirmatrelvir, All Descendants |
| 769518525 | budesonide |
| 883259145 | albuterol |
| 1000011148 | Dexamethasone--non otic, ophthalm, MDI |

| | |
|---|---|
| 1000033171 | N3C Lisinopril |
| 56320741 | aspirin |
| 745170047 | [CVDT] Carvedilol |
| 940495880 | [CVDT] Methylprednisolone |
| 65058498 | [Pitt N3C] Prednisone |
| 472540083 | apixaban |
| 1000040403 | Remdesivir, All Descendants |

**Supplementary Table 4: N3C codeset identifiers and concept set names for 15 drugs administered in the acute COVID window.**

| Codeset ID | Concept Set Name |
|---|---|
| 577359343 | echocardiography procedures |
| 30670147 | electrocardiography procedures |
| 920228667 | radiologic examination, chest |
| 209464212 | intravenous infusion, for therapy or prophylaxis |
| 996569760 | continuous positive airway pressure |
| 5668392 | other respiratory ventilation |
| 993408479 | Specimen collection for SARS-CoV-2 |
| 147612319 | therapeutic, prophylactic, or diagnostic injection |
| 970208606 | critical care |
| 764129526 | inhalation treatment with aerosol or nebulizer |
| 632643495 | intravenous infusion, hydration |

**Supplementary Table 5: N3C codeset identifiers and concept set names for 11 procedures administered in the acute COVID window.**

| Characteristic | Control<br>N = 32,238 | LD<br>N = 16,119 |
|---|---|---|
| **Clinical Factors** | | |
| Number of Visits | 54 (21, 121) | 89 (39, 182) |
| Days Observed | 1,945 (1,374, 2,241) | 1,945 (1,373, 2,241) |

| | | |
|---|---|---|
| BMI | 28 (23, 33) | 28 (22, 33) |
| COVID Positive | 11,328 (35%) | 5,590 (35%) |
| **Race** | | |
| White | 25,069 (78%) | 12,536 (78%) |
| Black or African American | 3,197 (9.9%) | 1,598 (9.9%) |
| Asian | 862 (2.7%) | 431 (2.7%) |
| American Indian or Alaska Native | 142 (0.4%) | 71 (0.4%) |
| Native Hawaiian or Other Pacific Islander | 108 (0.3%) | 54 (0.3%) |
| Other/Unknown | 2,860 (8.9%) | 1,429 (8.9%) |
| Age | 56 (33, 70) | 56 (34, 70) |
| **Sex** | | |
| Female | 17,358 (54%) | 8,679 (54%) |
| Male | 14,060 (44%) | 7,030 (44%) |
| Unknown | 820 (2.5%) | 410 (2.5%) |
| **Comorbidities** | | |
| Congestive Heart Failure | 4,277 (13%) | 3,786 (23%) |
| Cardiac Arrhythmias | 11,241 (35%) | 8,872 (55%) |
| Valvular Disease | 4,904 (15%) | 4,089 (25%) |
| Pulmonary Circulation Disorders | 2,438 (7.6%) | 2,544 (16%) |
| Peripheral Vascular Disorders | 4,208 (13%) | 3,524 (22%) |
| Uncomplicated Hypertension | 10,032 (31%) | 5,185 (32%) |
| Complicated Hypertension | 5,171 (16%) | 4,332 (27%) |
| Paralysis | 1,036 (3.2%) | 1,566 (9.7%) |
| Other Neurological Disorders | 5,040 (16%) | 5,764 (36%) |

| | | |
|---|---|---|
| Chronic Pulmonary Disease | 9,654 (30%) | 6,115 (38%) |
| Uncomplicated Diabetes | 2,286 (7.1%) | 1,343 (8.3%) |
| Complicated Diabetes | 5,177 (16%) | 4,375 (27%) |
| Hypothyroidism | 4,994 (15%) | 3,095 (19%) |
| Renal Failure | 4,301 (13%) | 3,797 (24%) |
| Liver Disease | 4,336 (13%) | 3,686 (23%) |
| Peptic Ulcer Disease without Bleeding | 902 (2.8%) | 851 (5.3%) |
| AIDS/HIV | 317 (1.0%) | 278 (1.7%) |
| Lymphoma | 477 (1.5%) | 355 (2.2%) |
| Metastatic Cancer | 1,421 (4.4%) | 1,351 (8.4%) |
| Solid Tumor without Metastasis | 2,826 (8.8%) | 1,538 (9.5%) |
| Rheumatoid Arthritis or Collagen Vascular Disease | 2,822 (8.8%) | 2,552 (16%) |
| Coagulopathy | 3,399 (11%) | 3,482 (22%) |
| Obesity | 8,884 (28%) | 5,550 (34%) |
| Weight Loss | 1,596 (5.0%) | 1,991 (12%) |
| Fluid and Electrolyte Disorders | 8,116 (25%) | 6,755 (42%) |
| Blood Loss Anemia | 923 (2.9%) | 871 (5.4%) |
| Deficiency Anemia | 3,557 (11%) | 3,103 (19%) |
| Alcohol Abuse | 5,094 (16%) | 5,103 (32%) |
| Drug Abuse | 3,220 (10.0%) | 3,044 (19%) |
| Psychoses | 1,026 (3.2%) | 845 (5.2%) |
| Depression | 9,768 (30%) | 6,683 (41%) |
| Elixhauser Score | 2 (-1, 15) | 11 (0, 29) |
| **Region** | | |
| Atlantic | 8,106 (25%) | 3,766 (23%) |
| Central | 15,616 (48%) | 8,045 (50%) |
| Southern | 2,394 (7.4%) | 1,248 (7.7%) |
| Western Pacific | 3,567 (11%) | 1,757 (11%) |
| Unknown | 2,555 (7.9%) | 1,303 (8.1%) |

**Supplementary Table 6: Characteristics of the Baseline Matched Cohort.** Summary table for patients in the Baseline Matched Cohort, including clinical factors, demographics (age, race, sex), comorbidities, and geographic region. Discrete data are displayed as “count (percentage)” and continuous data as “median (IQR)”.

| Lysosomal Disorder | N | Age | COVID Positive | Days Observed | Number of Visits | BMI | Elixhauser Score |
|---|---|---|---|---|---|---|---|
| Lipid storage disease | 6,543 | 65 (53, 75) | 2,391 (37%) | 1,926 (1,372, 2,201) | 83 (38, 161) | 29 (24, 34) | 8 (-1, 24) |
| Sphingolipidosis | 5,191 | 47 (27, 66) | 1,780 (34%) | 2,107 (1,575, 2,305) | 105 (43, 215) | 28 (22, 34) | 18 (5, 38) |
| Fabry's disease | 1,117 | 49 (33, 63) | 384 (34%) | 1,863 (1,296, 2,177) | 76 (31, 158) | 26 (21, 31) | 8 (0, 20) |
| Metachromatic leukodystrophy | 461 | 49 (21, 68) | 161 (35%) | 2,092 (1,433, 2,318) | 132 (57, 277) | 27 (19, 33) | 21 (10, 38) |
| Gaucher's disease | 446 | 47 (31, 68) | 151 (34%) | 1,553 (1,014, 2,049) | 56 (26, 134) | 24 (21, 29) | 6 (0, 19) |
| Mucopolysaccharidosis | 402 | 38 (18, 64) | 123 (31%) | 1,830 (1,149, 2,219) | 77 (34, 159) | 25 (20, 31) | 10 (0, 25) |
| MPS-I-H | 330 | 14 (7, 30) | 80 (24%) | 1,825 (1,150, 2,182) | 82 (40, 188) | 21 (17, 27) | 7 (0, 18) |
| Neuronal ceroid lipofuscinosis | 287 | 47 (18, 63) | 96 (33%) | 1,828 (1,170, 2,178) | 74 (38, 145) | 25 (20, 31) | 24 (10, 39) |
| Cystinosis | 228 | 30 (18, 46) | 76 (33%) | 1,907 (1,272, 2,235) | 117 (48, 254) | 23 (18, 28) | 13 (4, 27) |
| MPS-II | 215 | 30 (15, 63) | 72 (33%) | 1,890 (1,289, 2,216) | 90 (41, 178) | 25 (21, 31) | 6 (0, 24) |
| Tay-Sachs disease | 137 | 39 (24, 56) | 43 (31%) | 1,645 (888, 2,101) | 78 (33, 154) | 27 (21, 32) | 4 (-3, 19) |
| MPS-IV-A | 99 | 24 (12, 51) | 21 (21%) | 1,941 (1,458, 2,212) | 95 (42, 173) | 25 (21, 31) | 3 (0, 11) |
| Galactosylceramide beta-galactosidase deficiency | 94 | 23 (7, 38) | 25 (27%) | 1,517 (801, 1,935) | 57 (20, 105) | 22 (17, 28) | 10 (0, 17) |
| GM2 gangliosidosis | 72 | 76 (63, 83) | 31 (43%) | 2,027 (1,353, 2,228) | 114 (47, 210) | 26 (21, 30) | 18 (3, 33) |
| Niemann-Pick disease, type C | 66 | 28 (16, 40) | 21 (32%) | 1,729 (1,145, 2,106) | 82 (44, 144) | 22 (17, 27) | 13 (9, 30) |
| Gangliosidosis | 65 | 49 (6, 66) | 25 (38%) | 1,714 (935, 2,043) | 56 (33, 103) | 21 (18, 31) | 10 (0, 21) |

| | | | | | | | |
|---|---|---|---|---|---|---|---|
| Sphingomyelin/cholesterol lipidosis | 44 | 37 (30, 57) | < 20 (< 45%) | 1,561 (793, 2,089) | 66 (32, 83) | 28 (22, 33) | 2 (-2, 14) |
| Niemann-Pick disease, type B | 29 | 43 (29, 56) | < 20 (< 69%) | 1,364 (725, 2,010) | 64 (18, 172) | 25 (21, 30) | 13 (0, 27) |
| MPS-I-H/S | 29 | 21 (6, 30) | < 20 (< 69%) | 1,756 (879, 2,127) | 101 (55, 205) | 25 (20, 28) | 10 (0, 20) |
| Danon disease | 27 | 25 (20, 38) | < 20 (< 74%) | 1,756 (728, 2,254) | 144 (53, 218) | 25 (19, 34) | 20 (13, 42) |
| Cerebral lipidosis | 24 | 29 (9, 55) | < 20 (< 83%) | 1,775 (1,072, 2,165) | 119 (85, 156) | 21 (16, 27) | 10 (1, 20) |
| Sandhoff disease | 21 | 22 (6, 39) | < 20 (< 95%) | 1,656 (1,039, 2,034) | 74 (44, 131) | 23 (17, 29) | 8 (0, 18) |
| MPS-I-S | 21 | 30 (10, 46) | < 20 (< 95%) | 1,896 (1,288, 2,099) | 91 (46, 163) | 25 (19, 32) | 2 (0, 17) |
| Other | 171 | 22 (11, 47) | 42 (25%) | 1,600 (1,176, 2,160) | 87 (37, 190) | 21 (17, 27) | 10 (0, 21) |

**Supplementary Table 7: Lysosomal disorder characteristics in the Baseline Matched Cohort.**
Summary table for lysosomal disorder patients in the Baseline Matched Cohort, stratified by SNOMED lysosomal disorder subtype labels for increased granularity. “Other” contains subtypes with counts fewer than 20 patients. Discrete data are displayed as “count (percentage)” and continuous data as “median (IQR)”. Counts fewer than 20 are obscured with “< 20 (< X%)”.

| Elixhauser Comorbidity | Category | ICD-10 Prefixes |
|---|---|---|
| Congestive heart failure | Cardiometabolic | I099, I110, I130, I132, I255, I420, I425, I426, I427, I428, I429, I43, I50, P290 |
| Cardiac arrhythmias | Cardiometabolic | I441, I442, I443, I456, I459, I47, I48, I49, R000, R001, R008, T821, Z450, Z950 |
| Valvular disease | Cardiometabolic | A520, I05, I06, I07, I08, I091, I098, I34, I35, I36, I37, I38, I39, Q230, Q231, Q232, Q233, Z952, Z953, Z954 |
| Pulmonary circulation disorders | Cardiometabolic | I26, I27, I280, I288, I289 |
| Peripheral vascular disorders | Cardiometabolic | I70, I71, I731, I738, I739, I771, I790, I792, K551, K558, K559, Z958, Z959 |
| Hypertension, uncomplicated | Cardiometabolic | I10 |
| Hypertension, complicated | Cardiometabolic | I11, I12, I13, I15 |
| Diabetes, uncomplicated | Cardiometabolic | E100, E101, E109, E110, E111, E119, E120, E121, E129, E130, E131, E139, E140, E141, E149 |
| Diabetes, complicated | Cardiometabolic | E102, E103, E104, E105, E106, E107, E108, E112, E113, E114, E115, E116, E117, E118, E122, E123, E124, E125, E126, E127, E128, E132, E133, E134, E135, E136, E137, E138, E142, E143, E144, E145, E146, E147, E148 |
| Obesity | Cardiometabolic | E66 |
| Chronic pulmonary disease | Pulmonary | I278, I279, J40, J41, J42, J43, J44, J45, J46, J47, J60, J61, J62, J63, J64, J65, J66, J67, J684, J701, J703 |
| Renal failure | Renal | I120, I131, N18, N19, N250, Z490, Z491, Z492, Z940, Z992 |
| Liver disease | Hepatic/GI | B18, I85, I864, I982, K70, K711, K713, K714, K715, K717, K72, K73, K74, K760, K762, K763, K764, K765, K766, K767, K768, K769, Z944 |
| Peptic ulcer disease | Hepatic/GI | K257, K259, K267, K269, K277, K279, K287, K289 |
| Paralysis | Neurologic | G041, G114, G801, G802, G81, G82, G830, G831, G832, G833, G834, G839 |
| Other neurological disorders | Neurologic | G10, G11, G12, G13, G20, G21, G22, G254, G255, G312, G318, G319, G32, G35, G36, G37, G40, G41, G931, G934, R470, R56 |

| | | |
|---|---|---|
| Hypothyroidism | Endocrine | E00, E01, E02, E03, E890 |
| Coagulopathy | Hematologic | D65, D66, D67, D68, D691, D693, D694, D695, D696 |
| Blood loss anemia | Hematologic | D500 |
| Deficiency anemia | Hematologic | D508, D509, D51, D52, D53 |
| Weight loss | Nutritional | E40, E41, E42, E43, E44, E45, E46, R634, R64 |
| Fluid and electrolyte disorders | Nutritional | E222, E86, E87 |
| Lymphoma | Malignancy | C81, C82, C83, C84, C85, C88, C96, C900, C902 |
| Metastatic cancer | Malignancy | C77, C78, C79, C80 |
| Solid tumor, without metastasis | Malignancy | C00–C97 |
| Collagen vascular disease | Autoimmune | L940, L941, L943, M05, M06, M08, M120, M123, M30, M310, M311, M312, M313, M32, M33, M34, M35, M45, M461, M468, M469 |
| AIDS/HIV | Infectious | B20, B21, B22, B24 |
| Alcohol abuse | Substance Use | F10, E52, G621, I426, K292, K700, K703, K709, T51, Z502, Z714, Z721 |
| Drug abuse | Substance Use | F11, F12, F13, F14, F15, F16, F18, F19, Z715, Z722 |
| Psychoses | Psychiatric | F20, F22, F23, F24, F25, F28, F29, F302, F312, F315 |
| Depression | Psychiatric | F204, F313, F314, F315, F32, F33, F341, F412, F432 |

**Supplementary Table 8: Mapping of Elixhauser comorbidities to organ system categories and ICD-10 prefixes.**
Comorbidities were defined using ICD-10 code prefixes and were mapped to organ system categories.

| Severity | Median Δ | Mean Δ | Max Δ |
|---|---|---|---|
| Mild No ED | 7.95302 | 7.43935 | 20.1585 |
| Mild ED | 11.9213 | 11.9335 | 30.8186 |
| Moderate | 8.93014 | 8.82326 | 23.2213 |
| Severe | 6.93816 | 7.73853 | 19.0799 |
| Death | 6.00414 | 7.91564 | 19.9275 |

**Supplementary Table 9: Differences in comorbidity distributions across severity strata.**
Δ is defined as the difference in percentage prevalence between the LD and control groups (Δ = %LD − %control). Summary statistics (median, mean, and maximum Δ) are reported for each severity stratum.

| LD Category | OR (95% CI) | p-value | FDR q-value |
|---|---|---|---|
| Lysosomal disorder (Overall) | 0.98 (0.94, 1.02) | 0.287 | 0.788 |
| Sphingolipidosis | 0.95 (0.89, 1.01) | 0.082 | 0.637 |
| Lysosomal lipid storage disorder | 1.00 (0.96, 1.05) | 0.994 | 0.994 |
| Fabry disease | 1.02 (0.86, 1.20) | 0.822 | 0.994 |
| Gaucher disease | 1.12 (0.87, 1.45) | 0.364 | 0.800 |
| Metachromatic leukodystrophy | 0.85 (0.67, 1.08) | 0.174 | 0.637 |
| Inborn disorder of lysosomal amino acid transport | 0.95 (0.67, 1.34) | 0.771 | 0.994 |
| Gangliosidosis | 1.28 (0.93, 1.77) | 0.125 | 0.637 |
| Cystinosis | 0.98 (0.69, 1.38) | 0.907 | 0.994 |
| Neuronal ceroid lipofuscinosis | 1.02 (0.75, 1.39) | 0.875 | 0.994 |
| Glycoproteinosis | 0.91 (0.32, 2.60) | 0.860 | 0.994 |

**Supplementary Table 10: Conditional logistic regression for SARS-CoV-2 infection in the Baseline Matched Cohort by lysosomal disorder category.**
Odds ratios (OR) for SARS-CoV-2 infection. Models were conditioned on matching variables (age, sex, race, days observed, and data contributing site). "—" indicates estimates not estimated; mucopolysaccharidosis was not estimated due to dataset limitations.

| LD Category | Comorbidity Covariates |
|---|---|
| Sphingolipidosis | Cardiac arrhythmias, Hypertension (uncomplicated), Paralysis, Other neurological disorders, Diabetes (uncomplicated), Diabetes (complicated), Liver disease, Collagen vascular disease, Weight loss, Fluid and electrolyte disorders, Alcohol abuse |
| Lysosomal lipid storage disorder | Cardiac arrhythmias, Paralysis, Other neurological disorders, Diabetes (complicated), Liver disease, Collagen vascular disease, Weight loss, Deficiency anemia, Alcohol abuse |
| Fabry disease | Valvular disease, Renal failure |
| Gaucher disease | Coagulopathy |
| Metachromatic leukodystrophy | Paralysis, Other neurological disorders |
| Gangliosidosis | Other neurological disorders |
| Neuronal ceroid lipofuscinosis | Other neurological disorders |

**Supplementary Table 11: Comorbidity covariates included in adjusted models by lysosomal disorder (LD) category.**
Covariates were selected using category-specific logistic regression analyses ($p < 0.001$), regardless of the direction of association, and were included as covariates in adjusted conditional logistic regression models.

| Comparison | Group | N at risk | Events | Survival probability (95% CI) |
|---|---|---|---|---|
| Overall: LD vs Control | LD | 5,106 | 264 | 0.952 (0.946, 0.957) |
| Overall: LD vs Control | Control | 10,306 | 328 | 0.970 (0.967, 0.973) |
| LD: Hospitalized vs Non-Hospitalized | Hospitalized | 937 | 209 | 0.821 (0.799, 0.843) |
| LD: Hospitalized vs Non-Hospitalized | Non-Hospitalized | 4,169 | 55 | 0.987 (0.984, 0.991) |
| Control: Hospitalized vs Non-Hospitalized | Hospitalized | 1,161 | 264 | 0.820 (0.800, 0.840) |
| Control: Hospitalized vs Non-Hospitalized | Non-Hospitalized | 9,145 | 64 | 0.993 (0.992, 0.995) |

**Supplementary Table 12: One-year survival probabilities stratified by lysosomal disorder (LD) status and hospitalization.**
Survival probabilities are presented as Kaplan–Meier estimates with 95% confidence intervals (CI). N at risk and cumulative events are shown for each group.

#### Reporting Summary

**The RECORD statement – checklist of items, extended from the STROBE statement, that should be reported in observational studies using routinely collected health data.**

| | Item No. | STROBE items | Location in manuscript where items are reported | RECORD items | Location in manuscript where items are reported |
|---|---|---|---|---|---|
| **Title and abstract** | | | | | |
| | 1 | (a) Indicate the study's design with a commonly used term in the title or the abstract<br>(b) Provide in the abstract an informative and balanced summary of what was done and what was found | Abstract | RECORD 1.1: The type of data used should be specified in the title or abstract. When possible, the name of the databases used should be included.<br><br>RECORD 1.2: If applicable, the geographic region and timeframe within which the study took place should be reported in the title or abstract.<br><br>RECORD 1.3: If linkage between databases was conducted for the study, this should be clearly stated in the title or abstract. | Abstract ("retrospective cohort study… electronic health record data… National Clinical Cohort Collaborative (N3C)")<br><br>Abstract ("nationwide U.S…. longitudinal records starting January 1, 2018… captured between January 1, 2020, and July 11, 2024")<br><br>N/A (single centralized database, no linkage) |
| **Introduction** | | | | | |
| Background rationale | 2 | Explain the scientific background and rationale for the investigation being reported | Abstract; Background | | |
| Objectives | 3 | State specific objectives, including any prespecified hypotheses | Background (final paragraph) | | |
| **Methods** | | | | | |
| Study Design | 4 | Present key elements of study design early in the paper | Methods (Data Source & Preprocessing) | | |

| | | | | | |
|---|---|---|---|---|---|
| Setting | 5 | Describe the setting, locations, and relevant dates, including periods of recruitment, exposure, follow-up, and data collection | Methods (Data Source & Key Definitions) | | |
| Participants | 6 | *(a) Cohort study* - Give the eligibility criteria, and the sources and methods of selection of participants. Describe methods of follow-up<br><br>*(b) Cohort study* - For matched studies, give matching criteria and number of exposed and unexposed | | RECORD 6.1: The methods of study population selection (such as codes or algorithms used to identify subjects) should be listed in detail. If this is not possible, an explanation should be provided.<br><br>RECORD 6.2: Any validation studies of the codes or algorithms used to select the population should be referenced. If validation was conducted for this study and not published elsewhere, detailed methods and results should be provided.<br><br>RECORD 6.3: If the study involved linkage of databases, consider use of a flow diagram or other graphical display to demonstrate the data linkage process, including the number of individuals with linked data at each stage. | Methods (Key Definitions, Patient Identification and Classification, Survival analysis)<br><br>Methods (Patient Identification and Classification)<br><br>N/A (single centralized database, no linkage) |
| Variables | 7 | Clearly define all outcomes, exposures, predictors, potential confounders, and effect modifiers. Give diagnostic criteria, if applicable. | Methods (Key Definitions & Comorbidity Assessment) | RECORD 7.1: A complete list of codes and algorithms used to classify exposures, outcomes, confounders, and effect modifiers should be provided. If these cannot be reported, an explanation should be provided. | Methods (Patient Identification, Key Definitions, Comorbidity Assessment); Supplementary Tables 1, 4, 5, 8, and 11 |

| | | | | | |
|---|---|---|---|---|---|
| Data sources/ measurement | 8 | For each variable of interest, give sources of data and details of methods of assessment (measurement).<br>Describe comparability of assessment methods if there is more than one group | Methods (Data Source & Key Definitions) | | |
| Bias | 9 | Describe any efforts to address potential sources of bias | Methods (Data Preprocessing and Cohort Construction, Comorbidity Assessment, Survival analysis) | | |
| Study size | 10 | Explain how the study size was arrived at | Methods (Cohort Construction) | | |
| Quantitative variables | 11 | Explain how quantitative variables were handled in the analyses. If applicable, describe which groupings were chosen, and why | Methods (Data Preprocessing & Comorbidity Assessment) | | |
| Statistical methods | 12 | (a) Describe all statistical methods, including those used to control for confounding<br>(b) Describe any methods used to examine subgroups and interactions<br>(c) Explain how missing data were addressed<br>(d) *Cohort study* - If applicable, explain how loss to follow-up was addressed<br>(e) Describe any sensitivity analyses | Methods (Data Preprocessing and Cohort Construction & Statistical Analysis)<br><br><br>N/A<br><br>N/A | | |
| Data access and cleaning methods | | .. | | RECORD 12.1: Authors should describe the extent to which the investigators had access to the database population used to create the study population. | Methods (Data Source); Declarations (Availability of data and materials) |

| | | | | | |
|---|---|---|---|---|---|
| | | | | RECORD 12.2: Authors should provide information on the data cleaning methods used in the study. | Methods (Data Source - harmonization of 85 sites into OMOP CDM) |
| Linkage | | .. | | RECORD 12.3: State whether the study included person-level, institutional-level, or other data linkage across two or more databases. The methods of linkage and methods of linkage quality evaluation should be provided. | N/A (single centralized database, no linkage) |
| **Results** | | | | | |
| Participants | 13 | (a) Report the numbers of individuals at each stage of the study (*e.g.*, numbers potentially eligible, examined for eligibility, confirmed eligible, included in the study, completing follow-up, and analysed)<br>(b) Give reasons for non-participation at each stage.<br>(c) Consider use of a flow diagram | Results (first paragraph); Figure 1 & Supplementary Figure 1 | RECORD 13.1: Describe in detail the selection of the persons included in the study (*i.e.,* study population selection) including filtering based on data quality, data availability and linkage. The selection of included persons can be described in the text and/or by means of the study flow diagram. | Methods (Cohort Construction)<br>Results (first paragraph); Figure 1 & Supplementary Figure 1 |
| Descriptive data | 14 | (a) Give characteristics of study participants (*e.g.*, demographic, clinical, social) and information on exposures and potential confounders<br>(b) Indicate the number of participants with missing data for each variable of interest<br>(c) *Cohort study* - summarise follow-up time (*e.g.*, average and total amount) | Table 1; Supplementary Tables 6 and 7<br><br>Results (Matched cohort characteristics); Supplementary Figure 1<br>Table 1 | | |
| Outcome data | 15 | *Cohort study* - Report numbers of outcome events or summary measures over time | Table 1 (Severity Type); Supplementary Table 12 | | |

| | | | | | |
|---|---|---|---|---|---|
| Main results | 16 | (a) Give unadjusted estimates and, if applicable, confounder-adjusted estimates and their precision (e.g., 95% confidence interval). Make clear which confounders were adjusted for and why they were included<br>(b) Report category boundaries when continuous variables were categorized<br>(c) If relevant, consider translating estimates of relative risk into absolute risk for a meaningful time period | Table 2; Results (throughout)<br>N/A<br>N/A | | |
| Other analyses | 17 | Report other analyses done—e.g., analyses of subgroups and interactions, and sensitivity analyses | Results (Comorbidity stratification, survival, acute-phase interventions); Figures 2-4; Supplementary Figures 5-8 | | |
| **Discussion** | | | | | |
| Key results | 18 | Summarise key results with reference to study objectives | Conclusions | | |
| Limitations | 19 | Discuss limitations of the study, taking into account sources of potential bias or imprecision. Discuss both direction and magnitude of any potential bias | Discussion (paras. 7-9) | RECORD 19.1: Discuss the implications of using data that were not created or collected to answer the specific research question(s). Include discussion of misclassification bias, unmeasured confounding, missing data, and changing eligibility over time, as they pertain to the study being reported. | Discussion (paras. 7-9) |
| Interpretation | 20 | Give a cautious overall interpretation of results considering objectives, limitations, multiplicity of analyses, results from similar studies, and other relevant evidence | Discussion (throughout) | | |

| | | | | | |
|---|---|---|---|---|---|
| Generalisability | 21 | Discuss the generalisability (external validity) of the study results | Discussion (para. 10) | | |
| **Other Information** | | | | | |
| Funding | 22 | Give the source of funding and the role of the funders for the present study and, if applicable, for the original study on which the present article is based | Declarations (Funding) | | |
| Accessibility of protocol, raw data, and programming code | | .. | | RECORD 22.1: Authors should provide information on how to access any supplemental information such as the study protocol, raw data, or programming code. | Declarations (Availability of data and materials) |

*Reference: Benchimol EI, Smeeth L, Guttmann A, Harron K, Moher D, Petersen I, Sørensen HT, von Elm E, Langan SM, the RECORD Working Committee. The REporting of studies Conducted using Observational Routinely-collected health Data (RECORD) Statement. *PLoS Medicine* 2015; in press.

*Checklist is protected under Creative Commons Attribution (CC BY) license.
